# Walking in the Free World: Establishing Normative Trajectories for Ecological Assessment of Robust Gait Variability with Age

**DOI:** 10.64898/2026.03.06.26347806

**Authors:** Kai Zhe Tan, Krešimir Friganović, Yong Kuk Kim, Angela Frautschi, Michelle Gwerder, Kok Yang Tan, Vanessa Jean Wen Koh, Rahul Malhotra, Angelique Wei-Ming Chan, David B. Matchar, Navrag B. Singh

**Affiliations:** Future Health Technologies Programme, Singapore-ETH Centre, Singapore, Singapore; Built Environment in Falls & Arthritis Project, Singapore-ETH Centre, Singapore, Singapore; Institute for Biomechanics, ETH Zurich, Zurich, Switzerland; Programme in Health Services Research & Population Health (HSRPH), Duke-NUS Medical School, Singapore; Centre for Ageing Research & Education (CARE), Duke-NUS Medical School, Singapore

**Author notes:** Contributing authors.

**Keywords:** Gait variability, wearable sensors, ageing, fall risk

## Abstract

Gait variability is a critical functional indicator of dynamic balance and neurocognitive decline in health. Its translation into clinical practice is, however, challenged by a lack of age-related normative trajectories and reference values under real-world ecological settings. Furthermore, the conventional metrics used to estimate gait variability (Coefficient of Variation, **CV**; Standard Deviation, **SD**) have a fundamental methodological flaw: the inherent sensitivity of conventional metrics to the statistical outliers and environmental noise in real-world walking. In this study, we mitigate this factor by applying a robust statistical framework to quantify gait variability. Analysing a large-scale cohort of community-dwelling older adults (n=2,193), we first demonstrate that freeliving gait data follows a heavy-tailed distribution, necessitating the use of robust estimators like the Robust Coefficient of Variation (**RCV**_**MAD**_) and Median Absolute Deviation (**MAD**). Leveraging these metrics, we established the normative trajectory and reference values of real-world gait variability across the ageing lifespan, revealing a distinct, age-dependent increase in spatio-temporal fluctuations, indicating a decline in rhythmicity and steadiness with age. We further demonstrated the clinical utility of these robust metrics: **RCV**_**MAD**_ consistently yielded larger effect sizes than conventional **CV** in discriminating between fallers and non-fallers across all gait parameters. Furthermore, we illustrate the potential of long-term unsupervised monitoring to capture intrinsic variability during real-world walking. Validated for consistency and reliability, this robust framework provides the necessary ecological validity to transform gait variability into a standardised, rapid clinical metric for assessing functional decline at an early timepoint.

## 1 Introduction

Measurement of fluctuation in physiological signals, such as heart rate variability, is one of the fundamental components of data-driven medical diagnostics, providing quantitative indicators into human autonomic functions and overall health status [1– 3]. In movement science, gait variability is the natural spatio-temporal fluctuations during walking from one step to the next step. Far from being random noise, increased gait variability is a quantifiable biomarker of compromised dynamic balance [4, 5] and a promising indicator of early neurocognitive decline [6]. The proliferation of wearable Inertial Measurement Units (IMUs) [7–10] allows transition from laboratory to real-world monitoring and strengthens the uptake of gait variability as a viable digital biomarker for functional status. However, real-world walking typically involves transients such as gait initiation, termination, turns, pauses, and obstacle navigation that create statistical outliers. These anomalies artificially inflating the Coefficient of Variation (CV) and Standard Deviation (SD) — the conventional gold-standard metrics for quantifying gait variability [5], exposing their fundamental weakness [11] and rendering them unreliable [7, 12–14]. To address this, robust statistics offer alternative estimators (e.g., Median Absolute Deviation, MAD and Interquartile Range, IQR) that are inherently less sensitive to environmental artifacts [11], and therefore allow precise quantification of intrinsic biological rhythmicity even in “messy” real-world data.

Recent studies have shown that walking environment and surface characteristics contaminate gait parameters and adversely impact CV and SD of gait parameters [15, 16]. While robust variability metrics have shown promise in simulated datasets and controlled experiments [17], their validity in complex, free-living environments remains under-explored. Prior to clinical adoption, there is a critical need for rigorous validation under ecological valid real-world environment. While cut-off values and recommendations on testing protocols have allowed widespread acceptance of walking speed as an indicator of functional capacity [18, 19], such standardisation is currently missing for gait variability. Systematic reviews have consistently established that gait variability increases with age, reflecting a progressive physiological decline in dynamic balance [20]. However, in the absence of normative trajectories of these robust metrics across the older adult lifespan, clinicians remain unable to reliably distinguish expected age-related changes from pathological deviations. It is crucial to establish ecologically valid, age-stratified normative values for robust gait variability to serve as a reference standard for the aging population.

Gait variability has emerged as a promising digital biomarker for predicting falls early [4, 20, 21], and tracking progression of neurological and musculoskeletal disorders, including Parkinson’s disease [22–24]. However, the predictive power of conventional metrics is often affected by the measurement noise in daily living conditions. By applying robust metrics that filter extrinsic noise, we can re-evaluate the true diagnostic utility of gait variability for discriminating fall risk in large-scale community-dwelling older adults population cohorts. Furthermore, these metrics facilitate reliable long-term unsupervised monitoring, enabling the unobtrusive assessment of gait variability and detection of subtle longitudinal changes in gait quality — potentially enhancing diagnosis fidelity during sporadic clinical assessments.

To bridge the gap between biological insight and clinical application, the primary aim of this study was to define the normative trajectory of real-world robust gait variability, establishing robust, ecologically valid reference standards that account for the natural heterogeneity of aging. Second, we evaluated the diagnostic utility of these robust metrics by assessing their sensitivity to fall risk in community-dwelling older adults and their applicability for long-term unsupervised monitoring. Finally, to validate the methodological foundation of these insights, we systematically compared the intra-subject consistency and reliability of robust statistical measures (i.e., RCV_MAD_, RCV_IQR_, MAD, IQR) against conventional variability metrics (i.e., CV, SD) during treadmill walking and continuous free-living overground walking (Figure 1). By establishing statistically robust and ecologically valid benchmarks, this study enhances the precision of wearable sensors for assessing mobility, fall risk, and frailty in the real world and thereby improving the uptake of gait variability as biomarker for real-world monitoring of functional status.

**Fig. 1.**
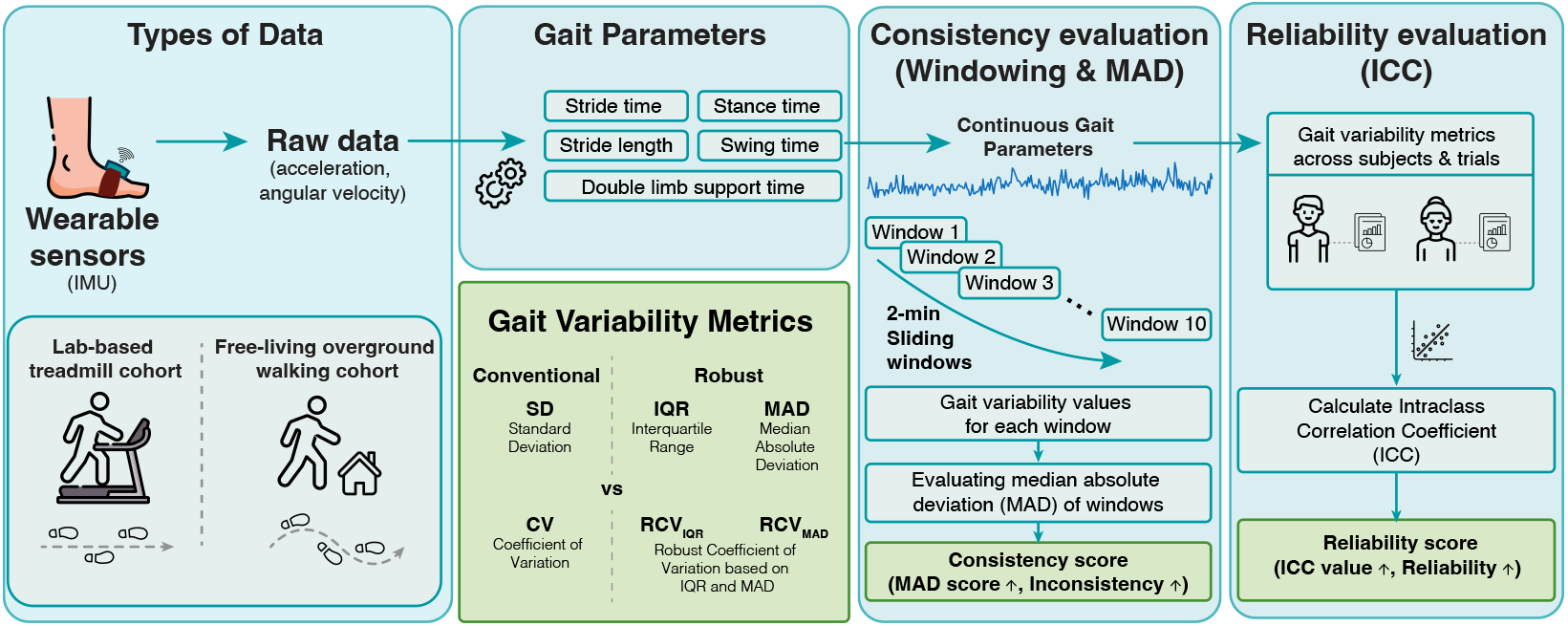
Schematic showcase our technical validation and evaluation framework for estimating consistency and reliability under lab-based settings and real-world condition.

## 2 Results

The results of this study are presented in five main sections, structured to first define the biological phenotype of aging mobility and then demonstrate the technical and clinical superiority of robust metrics in capturing it. First, we characterise the fundamental distributional properties of real-world gait parameters, identifying the heavy-tailed nature of free-living data that necessitates robust statistical frameworks. Second, leveraging these robust methods, we establish normative age-related trajectories and quantify the rate of movement rhythmicity decline across the ageing lifespan. Third, we evaluate the clinical utility of these gait variability metrics by assessing the ability to discriminate between community-dwelling older adults with and without history of falls (Case Study 1). Fourth, we illustrate the practical implications of gait variability metrics during long-term unsupervised monitoring of walking bouts in the community (Case Study 2). Finally, to underpin these biological and clinical findings, we provide a comprehensive technical validation, comparing the intra-subject consistency and reliability of robust versus conventional variability measures across two distinct cohorts of older adults: one with controlled treadmill walking in a laboratory setting and another performing free-living, real-world walking (referred to as overground walking).

### 2.1 Real-world mobility data is inherently heavy-tailed

Prior to gait variability analysis, we inspected the statistical distribution properties of the gait parameters for each participants. Contrary to the assumption of normality often implied in computing standard deviation, the data frequently exhibited deviations from normality, possessing both asymmetry (skewness) and heavy tails (kurtosis). Notably, temporal (i.e. stride time, stance time etc.) and spatial parameter (i.e. stride length) skewed in opposite directions in the overground data — temporal parameters towards longer durations (positive skew) and spatial parameters towards shorter lengths (negative skew).

The summary of statistical distribution for both cohorts is presented in Table 1. While treadmill data was moderately heavy-tailed (median kurtosis of 3.6), skewness was negligible across spatial and temporal parameters. In sharp contrast, the transition to overground walking resulted in a severe deviation from normality. The median kurtosis nearly doubled with high skewness in both spatial and temporal parameters with 60.3% of gait parameters exhibiting extreme kurtosis (*>* 6.0). This result indicates high presence of outliers and asymmetric stride distributions in overground walking data that violate the assumptions of normality required for reliable computations of the standard deviation.

**Table 1.**
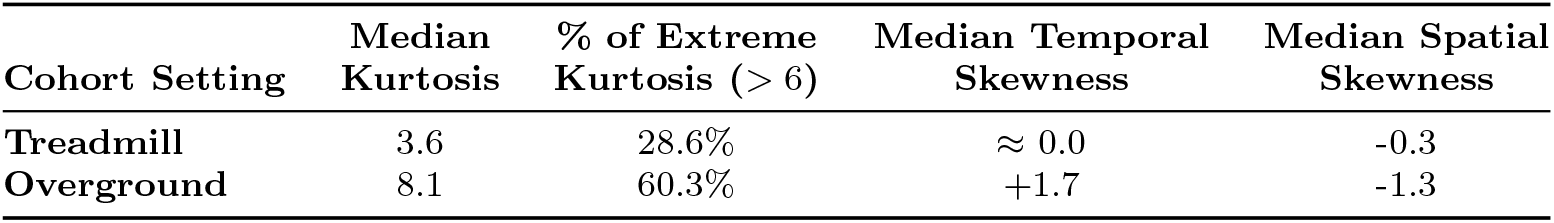
Statistical distribution characteristics of gait parameters across cohorts (perfect normal distributed has a kurtosis of 3.0 and skewness of 0).

### 2.2 Normative trajectories reveal a pervasive age-dependent loss of ambulatory rhythmicity

To identify the true phenotype of aging mobility masked by environmental noise, we applied Generalised Additive Models for Location, Scale, and Shape (GAMLSS) to our large-scale free-living overground walking cohort using robust variability metric RCV_MAD_. This framework allowed us to construct centile reference charts for real-world gait variability, accounting for the heavy-tailed distributions identified in our initial distribution analysis.

We found significant and systematic increase in robust gait variability across the adult lifespan. The main effect of age was significant (p*<*0.001) for nearly all evaluated spatio-temporal parameters (Appendix A: Table A1), confirming that the robust variability metric RCV_MAD_ is a sensitive biomarker of biological aging. The magnitude of this functional decline was clinically substantial. We quantified the “rate of aging” as the median percentage point increase in variability per decade. Spatial consistency showed the most profound deterioration, with Left Stride Length Variability increasing by approximately 1.29 to 1.45 percentage points per decade (Females: 1.29 [95% confidence interval: 0.98–1.60]; Males: 1.45 [95% confidence interval: 1.03–1.86]). This magnitude of decline is substantial, representing an approximate 10–20% relative worsening of spatial gait variability for every ten years of life. Similarly, Left Swing Time Variability worsened by 0.78 (Females) and 0.72 (Males) percentage points per decade. These quantitative estimates provide benchmarks for the expected rate of mobility decline in real-world stability. However, interaction testing revealed no statistically significant difference in the rate of age-related decline between males and females for any variability parameter (p*>*0.05 for all Age and Sex interactions; Appendix A: Table A1). This suggests that the progressive depletion of ambulatory rhythmicity reflects a fundamental, conserved biological process of aging that operates independently of sex-specific factors in this cohort.

Based on these robust models, we established sex-specific normative reference standards for free-living gait variability (Figure 2). These centile charts (3rd, 15th, 50th, 85th, and 97th percentiles) show the normative values for community-dwelling adults aged 60 to 90 years. The trajectories for key parameters, including Stride Time and Stride Length Variability, illustrate a continuous increase in walking variability, providing a reference framework to distinguish expected biological aging from pathological deviation in clinical monitoring (Figure A1 in Appendix A provides the comprehensive plots for all 12 parameters). Comprehensive age- and sex-stratified percentile lookup values for all robust parameters are provided as a standalone PDF in Supplementary Data 1.

**Fig. 2.**
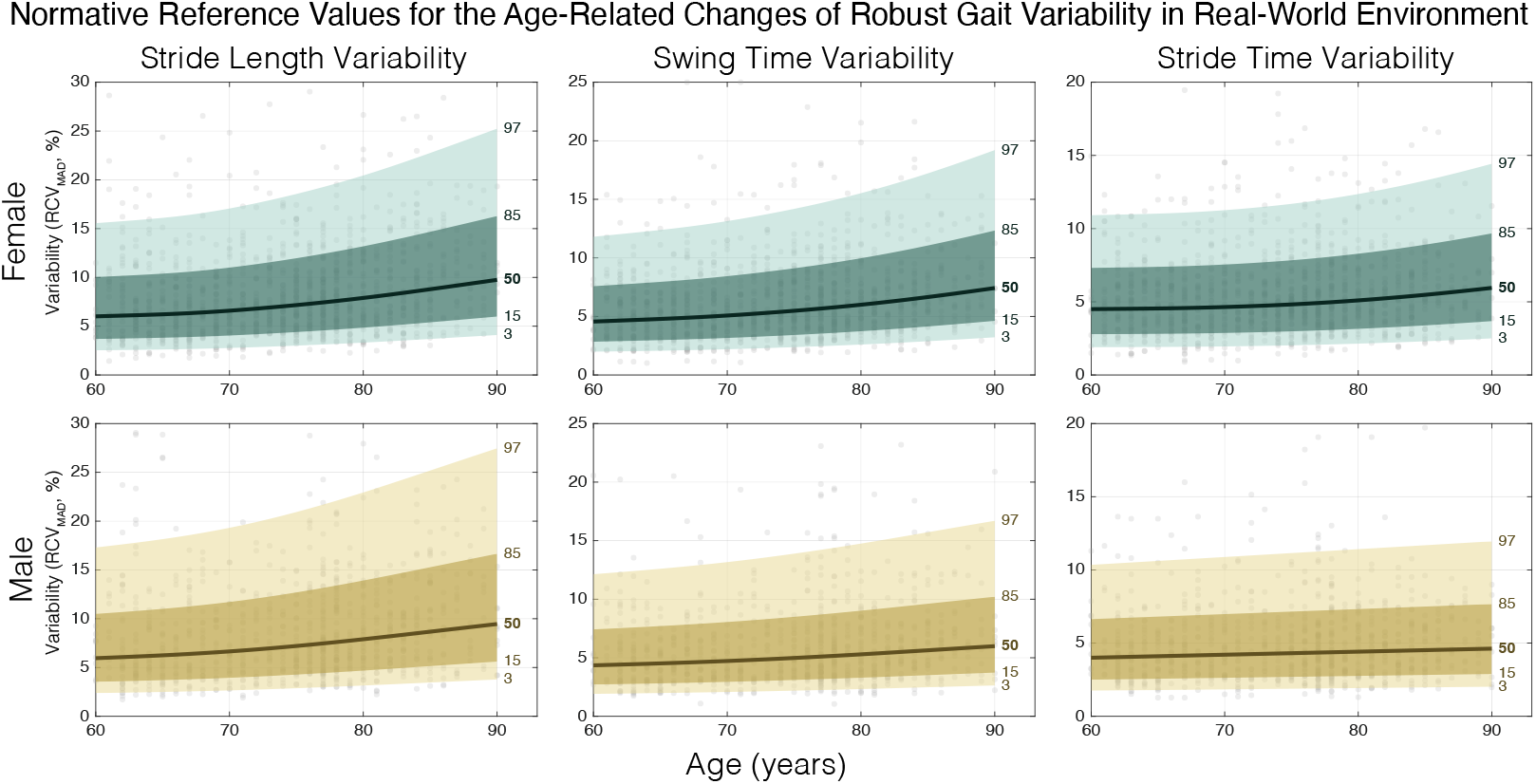
Age-related trajectories of robust gait variability (RCV_MAD_) in free-living environments: Generalised Additive Models for Location, Scale, and Shape (GAMLSS) centile charts (3rd, 15th, 50th, 85th, and 97th percentiles) depicting the normative changes of (a) Stride Length Variability, (b) Swing Time Variability, and (c) Stride Time Variability across the older adult lifespan (60–90 years), stratified by sex (Females: Green; Males: Yellow). The median (50th percentile) trajectory highlights a progressive loss of movement spatio-temporal rhythmicity, identifying robust variability as a sensitive digital biomarker for monitoring age-related functional decline.

### 2.3 Robust variability metrics enhances the discrimination of fall history in community-dwelling older adults

To assess and compare the clinical utility of the conventional and robust variability measures (CV and RCV_MAD_, respectively), we assessed their capacity to differentiate between individuals with and without a self-reported fall in the past year (Table 2). Both conventional and robust metrics identified statistically significant differences between the faller and non-faller groups for several key gait parameters. For instance, using the conventional CV, Right Swing Time was significantly higher in fallers (p = 0.006), as was Right Step Time (p = 0.011). On the other hand, the RCV_MAD_ identified significantly higher gait variability in fallers across almost all gait parameters, with the group difference in Right Swing Time being highly significant.

**Table 2.**
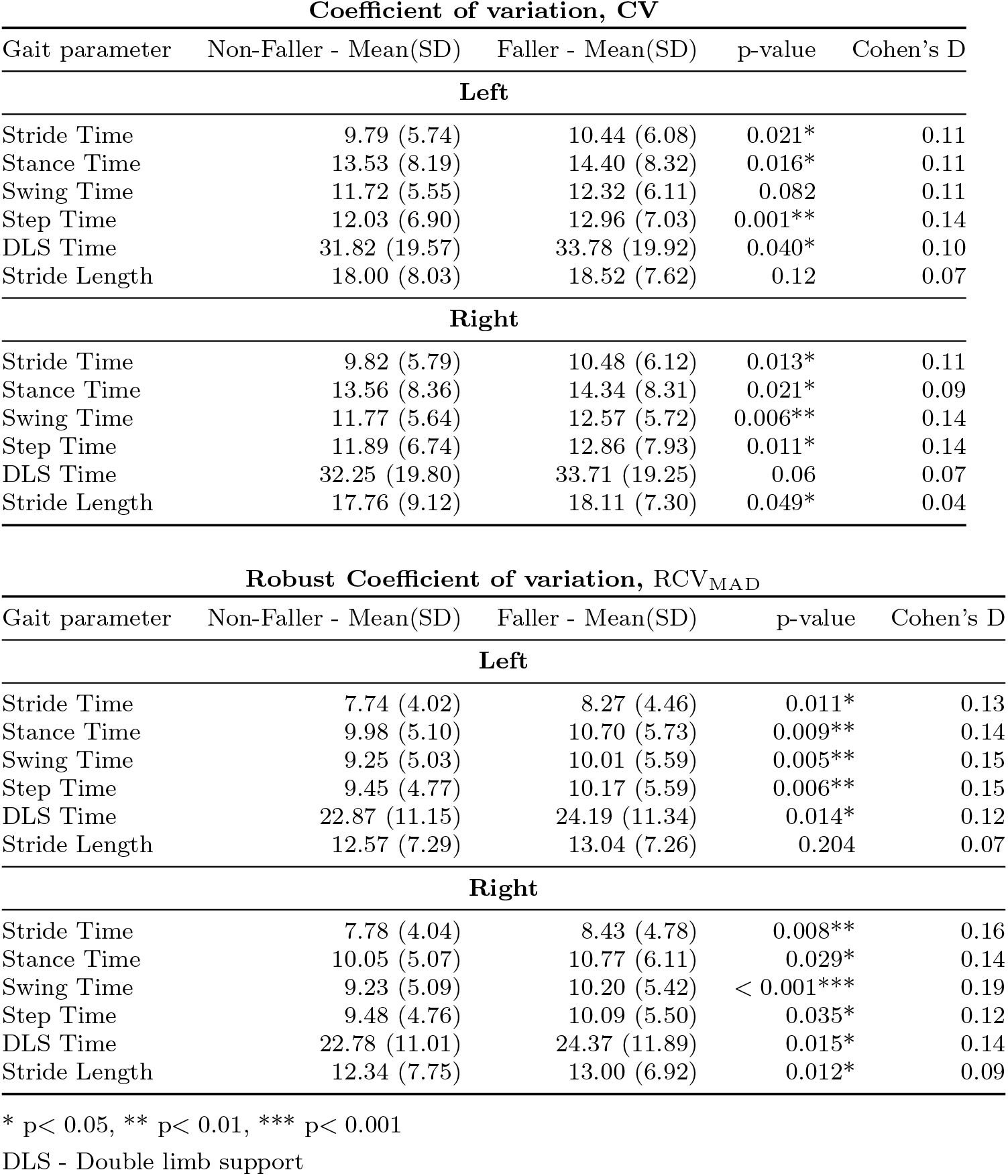
Comparison of gait variability metrics (CV and RCV_MAD_) between fallers and non-fallers.

| **Coefficient of variation, CV** |  |  |  |  |
| --- | --- | --- | --- | --- |
| Gait parameter | Non-Faller - Mean(SD) | Faller - Mean(SD) | p-value | Cohen's D |
| **Left** |  |  |  |  |
| Stride Time | 9.79 (5.74) | 10.44 (6.08) | 0.021* | 0.11 |
| Stance Time | 13.53 (8.19) | 14.40 (8.32) | 0.016* | 0.11 |
| Swing Time | 11.72 (5.55) | 12.32 (6.11) | 0.082 | 0.11 |
| Step Time | 12.03 (6.90) | 12.96 (7.03) | 0.001** | 0.14 |
| DLS Time | 31.82 (19.57) | 33.78 (19.92) | 0.040* | 0.10 |
| Stride Length | 18.00 (8.03) | 18.52 (7.62) | 0.12 | 0.07 |
| **Right** |  |  |  |  |
| Stride Time | 9.82 (5.79) | 10.48 (6.12) | 0.013* | 0.11 |
| Stance Time | 13.56 (8.36) | 14.34 (8.31) | 0.021* | 0.09 |
| Swing Time | 11.77 (5.64) | 12.57 (5.72) | 0.006** | 0.14 |
| Step Time | 11.89 (6.74) | 12.86 (7.93) | 0.011* | 0.14 |
| DLS Time | 32.25 (19.80) | 33.71 (19.25) | 0.06 | 0.07 |
| Stride Length | 17.76 (9.12) | 18.11 (7.30) | 0.049* | 0.04 |
| **Robust Coefficient of variation, $RCV_{MAD}$** |  |  |  |  |
| Gait parameter | Non-Faller - Mean(SD) | Faller - Mean(SD) | p-value | Cohen's D |
| **Left** |  |  |  |  |
| Stride Time | 7.74 (4.02) | 8.27 (4.46) | 0.011* | 0.13 |
| Stance Time | 9.98 (5.10) | 10.70 (5.73) | 0.009** | 0.14 |
| Swing Time | 9.25 (5.03) | 10.01 (5.59) | 0.005** | 0.15 |
| Step Time | 9.45 (4.77) | 10.17 (5.59) | 0.006** | 0.15 |
| DLS Time | 22.87 (11.15) | 24.19 (11.34) | 0.014* | 0.12 |
| Stride Length | 12.57 (7.29) | 13.04 (7.26) | 0.204 | 0.07 |
| **Right** |  |  |  |  |
| Stride Time | 7.78 (4.04) | 8.43 (4.78) | 0.008** | 0.16 |
| Stance Time | 10.05 (5.07) | 10.77 (6.11) | 0.029* | 0.14 |
| Swing Time | 9.23 (5.09) | 10.20 (5.42) | < 0.001*** | 0.19 |
| Step Time | 9.48 (4.76) | 10.09 (5.50) | 0.035* | 0.12 |
| DLS Time | 22.78 (11.01) | 24.37 (11.89) | 0.015* | 0.14 |
| Stride Length | 12.34 (7.75) | 13.00 (6.92) | 0.012* | 0.09 |

\* $p < 0.05$ , \*\* $p < 0.01$ , \*\*\* $p < 0.001$
DLS - Double limb support

An evaluation of Cohen’s D effect sizes confirmed that the RCV_MAD_ provides enhanced sensitivity. While the effect sizes for differentiating fallers were generally small across all metrics, the RCV_MAD_ consistently produced larger effect sizes than the CV for the same gait parameter. For example, the effect size for Right Swing Time increased from 0.14 when using CV to 0.19 when using RCV_MAD_. Similarly, the effect size for Right Stride Time increased from 0.11 to 0.16 using RCV_MAD_, a magnification of up to 45% in effect sizes. Although modest, this consistent amplification of the between-group difference suggests that RCV_MAD_ may offer improved sensitivity for detecting subtle gait alterations associated with fall risk, a critical feature for clinical screening and digital biomarker development.

### 2.4 Unsupervised monitoring reveals longitudinal patterns of intrinsic gait stability

To further illustrate the practical implications of gait variability metric choice in unsupervised, free-living bouts, Figure 3 shows a representative spatial distribution of gait variability of a outdoor walking bout, complemented with boxplots comparing the consistency of CV, RCV_IQR_ and RCV_MAD_ during 5 minute walking bouts segmented from long term monitoring.

**Fig. 3.**
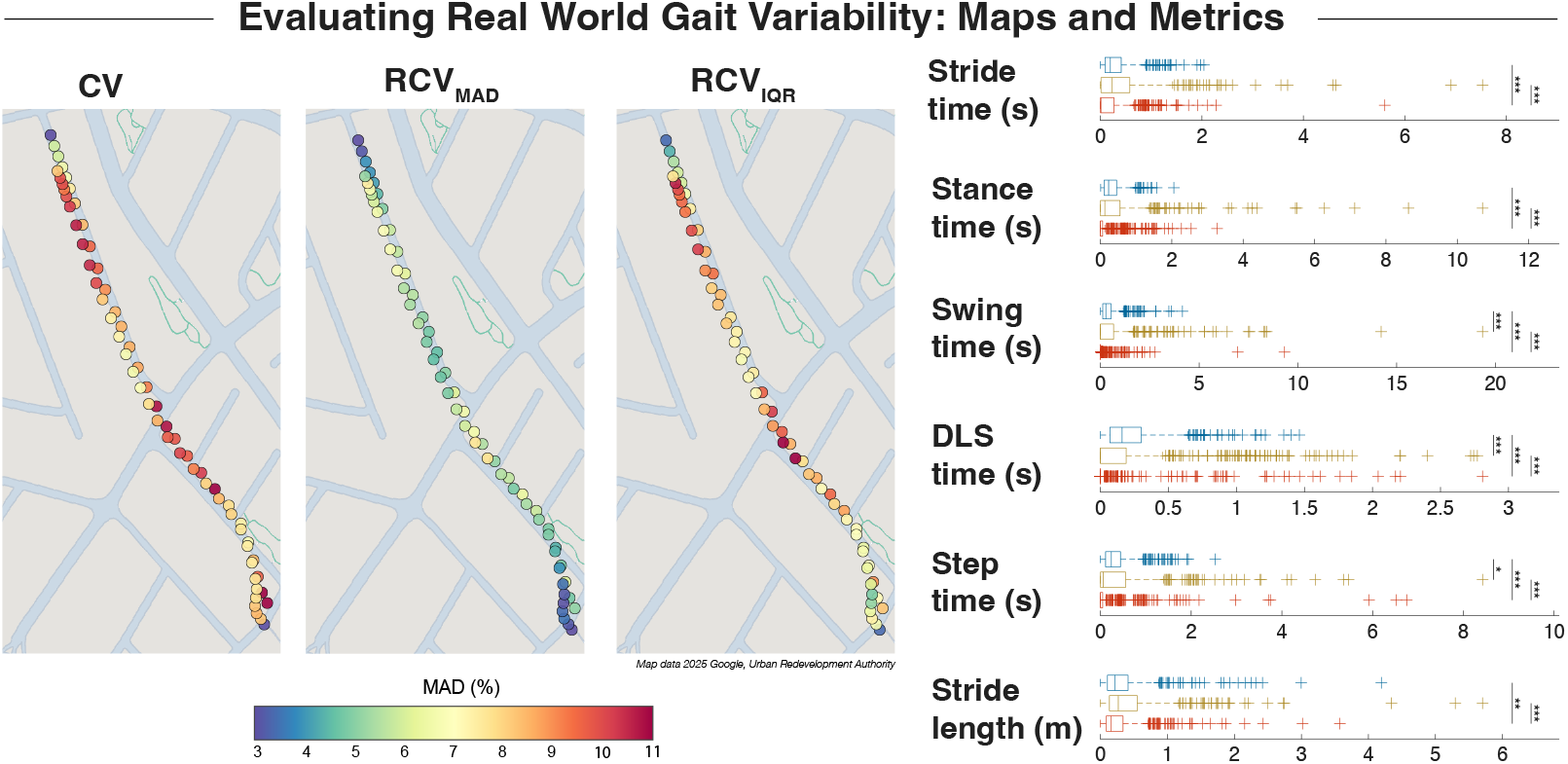
(Left) Representative GPS trace of a single long walking bout, color-coded by the magnitude of conventional CV, RCV_IQR_, and RCV_MAD_. The maps illustrate the spatial distribution of variability and highlight the susceptibility of CV and RCV_MAD_ to transient noise spikes. In comparison, RCV_MAD_ shows more uniformity. (Right) Boxplots comparing consistency across six gait parameters (left and right combined) of the 5 minute walking bouts during unsupervised longterm monitoring. Consistency is quantified as the MAD of ten 2-minute moving windows (lower y-axis values indicate higher consistency). Brackets indicate one-tailed Wilcoxon signed-rank tests testing the hypothesis sequence of decreasing inconsistency: CV *>* RCV_IQR_ *>* RCV_MAD_ (*p*<* 0.05, **p*<* 0.01, ***p*<* 0.001). The RCV_MAD_ demonstrated significantly higher consistency than CV and RCV_IQR_ across all conditions.

The map generated using the conventional CV and RCV_IQR_ exhibits a highly ‘noisy’ pattern, characterised by transient spikes of extreme variability along otherwise consistent straight paths. These spikes likely reflect the metric’s sensitivity to isolated non-steady-state walking steps or inherent sensor noise in unsupervised data. In contrast, the map based on RCV_MAD_ provides noticeably smoother and more continuous transitions in gait variability of the walking bout. Hotspots with higher variability in the RCV_MAD_ map appear less sporadic and potentially corresponding to actual environmental challenges (e.g. junctions, turns) rather than measurement noise. These results suggest that the conventional CV may overestimate physiological inconsistency in real-world monitoring due to its lack of robustness. Boxplots show results consistent with the treadmill and overground cohorts, where intra-subject consistency was statistically significantly higher with RCV_MAD_ compared to both RCV_IQR_ and CV.

### 2.5 Technical Validation of Gait Variability Metrics

#### 2.5.1 Consistency of gait variability metrics

To quantify the absolute intra-subject consistency of each metric, the MAD was calculated from the 10 windowed values of gait variability for each participant. This calculation provides a single “inconsistency score” for each metric, each parameter and each participant, where a lower value indicates higher consistency (i.e. less fluctuation across the windows). The distributions of these inconsistency scores for all participants are visualised using box plots in Figure 4 and 5. Statistical comparisons between metrics were performed using one-tailed Wilcoxon signed-rank tests.

**Fig. 4.**
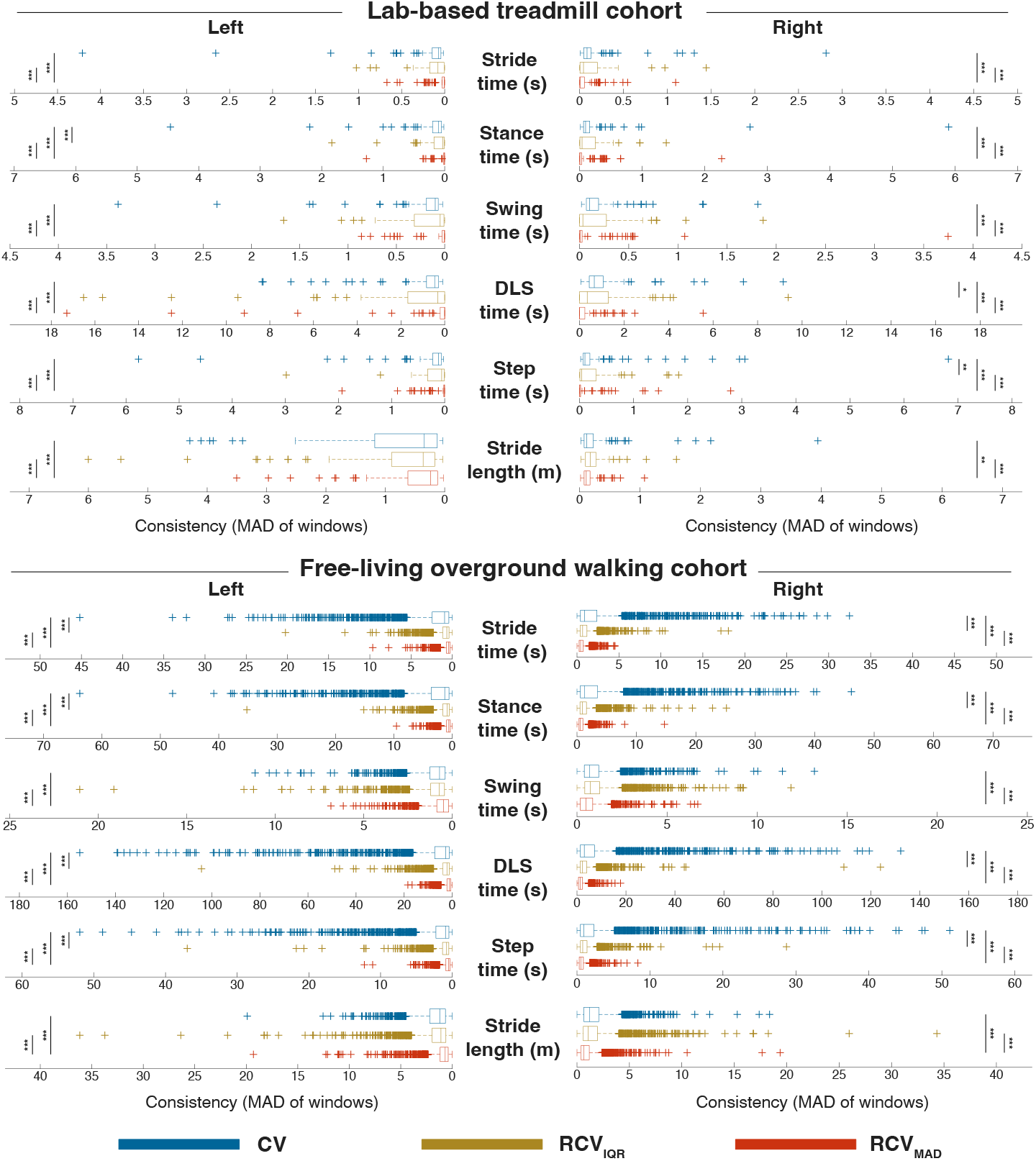
Boxplots comparing the intra-subject consistency of the conventional Coefficient of Variation (CV), the Robust CV based on the Interquartile Range (RCV_IQR_), and the Robust CV based on the Median Absolute Deviation (RCV_MAD_). Consistency is quantified by the Median Absolute Deviation (MAD) of the values calculated from ten 2-minute moving windows for each participant. Lower MAD values on the y-axis indicate higher consistency (less fluctuation). Data are shown for six gait parameters, separated by the left and right limbs, in two distinct cohorts: (top) lab-based treadmill cohort, (bottom) free-living overground walking cohort. Brackets indicate paired comparisons performed using one-tailed Wilcoxon signed-rank tests based on the hypotheses that 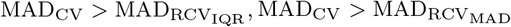 and 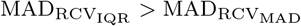. Asterisks denote the level of statistical significance: * p*<* 0.05, ** p*<* 0.01, *** p*<* 0.001. Across all conditions and parameters, RCV_MAD_ demonstrated significantly higher consistency than the traditional CV and RCV_IQR_.

**Fig. 5.**
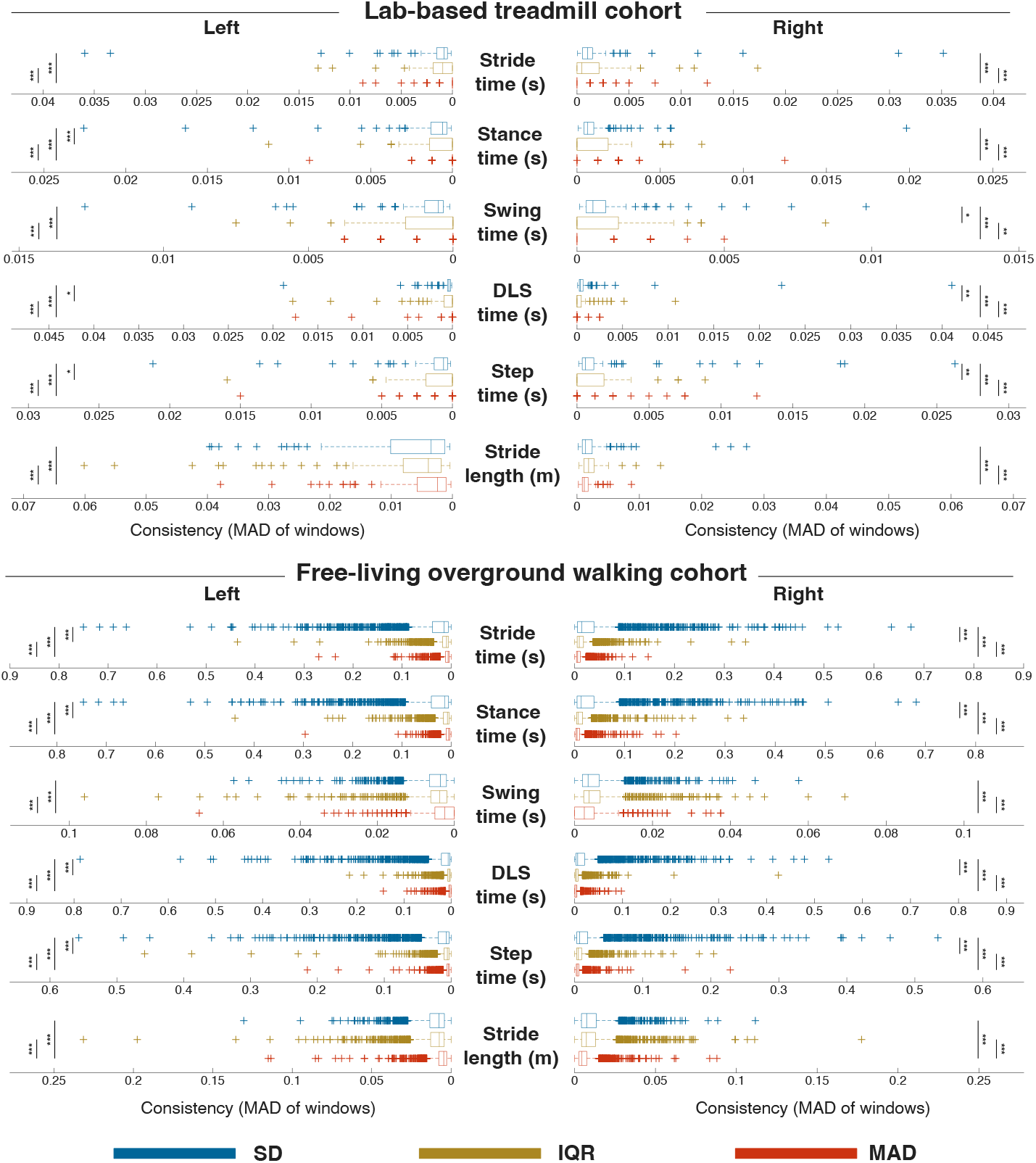
Boxplots comparing the intra-subject consistency of the Standard Deviation (SD), the Interquartile Range (IQR), and the Median Absolute Deviation (MAD). Consistency is quantified by the Median Absolute Deviation (MAD) of the values calculated from ten 2-minute moving windows for each participant. Lower MAD values on the y-axis indicate higher consistency (less fluctuation). Data are shown for six gait parameters, separated by the left and right limbs, in two distinct cohorts: (top) lab-based treadmill cohort, (bottom) free-living overground walking cohort. Brackets indicate paired comparisons performed using one-tailed Wilcoxon signed-rank tests based on the hypotheses that SD *>* IQR, SD *>* MAD and IQR *>* MAD. Asterisks denote the level of statistical significance: * p*<* 0.05, ** p*<* 0.01, *** p*<* 0.001. Across all conditions and parameters, MAD demonstrated significantly higher consistency than the traditional SD and IQR.

##### 2.5.1.1 *Consistency of* RCV_MAD_, RCV_IQR_ *and CV*

Among the three mean/median-adjusted relative dispersion measures, the RCV_MAD_ demonstrated noticeably better measurement consistency compared to the conventional CV. Across both laboratory and free-living walking conditions, RCV_MAD_ yielded significantly lower inconsistency scores for every gait parameter analysed (Figure 4, Table A2 in Appendix A). This improvement was statistically significant across all gait parameters (Wilcoxon signed-rank test, p *<* 0.001). For example, in the lab-based treadmill cohort, the median inconsistency for Left Swing Time CV was 0.098, whereas the corresponding value using RCV_MAD_ was 0.006, a 94% increase in consistency. Similar results were observed in the overground cohort, where the median inconsistency for Left Stride Time CV was 0.929, which dropped to 0.428 when using RCV_MAD_, a 54% increase in consistency.

On the other hand, the RCV_IQR_ produced inconsistent results. In lab-based treadmill data, RCV_IQR_ showed a median inconsistency similar to the conventional CV across most parameters, but with a much larger interquartile range, indicating poorer measure consistency. RCV_IQR_ was significantly more consistent than CV for most parameters in the overground cohort, with the exception of swing time and stride length.

The effect size for the improvement using RCV_MAD_ was further illustrated using the rank-biserial correlation (Table A2 in Appendix A). The effect sizes for the improvements of RCV_MAD_ over CV were consistently moderate to large, with ranges of 0.30 to 0.72 in treadmill and 0.42 to 0.67 in overground cohort. There is a strong evidence to support that RCV_MAD_ provides a more consistent characterisation of an individual’s intrinsic gait variability. In contrast, the RCV_IQR_ produced lower effect sizes than RCV_MAD_, occasionally showing lower consistency than the standard CV, suggesting it is a less consistent metric.

##### 2.5.1.2 Consistency of SD, IQR and MAD

Among the three absolute metrics of statistical dispersion used to quantify variability — SD, IQR, and MAD — MAD provided the most consistent measurements (Figure 5, Table A3 in Appendix A). Similarly to the RCV_MAD_ results, MAD invariably produced lower inconsistency scores than SD and IQR for all gait parameters in both cohorts. For example, in the overground cohort, the median inconsistency for Left Stance Time when measured with SD was 0.013, which was reduced by more than half to 0.005 when using MAD. Similarly, in the lab-based cohort, the inconsistency of Left Stride Length SD was 0.003, while the corresponding MAD value was 33% lower.

In contrast, the IQR was found to be an inconsistent metric for variability assessment, often yielding inconsistent results. In the lab-based cohort, the consistency of IQR was erratic, failing to show a consistent advantage over the less robust SD. IQR showed better consistency than CV for all gait parameters except swing time and stride length in the overground cohort, but not necessarily better than MAD. For instance, the median inconsistency score for Left Swing Time for the IQR was 5.3 % higher compared to SD and 47.6% higher compared to MAD, suggesting IQR lacks the required consistency for robust gait variability assessment.

The Wilcoxon signed-rank test confirmed that MAD was significantly more consistent than SD (p *<* 0.001) for all parameters. The effect size for the improvement of MAD over SD ranged from 0.29 to 0.72 in the treadmill cohort and from 0.42 to 0.68 in the overground cohort. These results strongly support the conclusion that MAD is a more robust and consistent metric for assessing gait variability compared to both SD and IQR.

### 2.5.2 Reliability of gait variability metrics

To assess the reliability of metrics, intraclass correlation coefficient (ICC) is calculated for each metric across the 10 walking windows. The ICC(A,1) values and their 95% confidence intervals of each parameter are visualised with plots, along with mean ICC for each metric, shown in Figure 6 and 7. The full results are presented in Table A4 (mean-/median-adjusted variability measure) and A5 (absolute variability measure), in Appendix A. ICC was interpreted using the guidelines established [25]: ‘poor’(*<* 0.50), ‘moderate (0.50–0.75)’, ‘good (0.75–0.90)’ and ‘excellent’ (*>* 0.90).

**Fig. 6.**
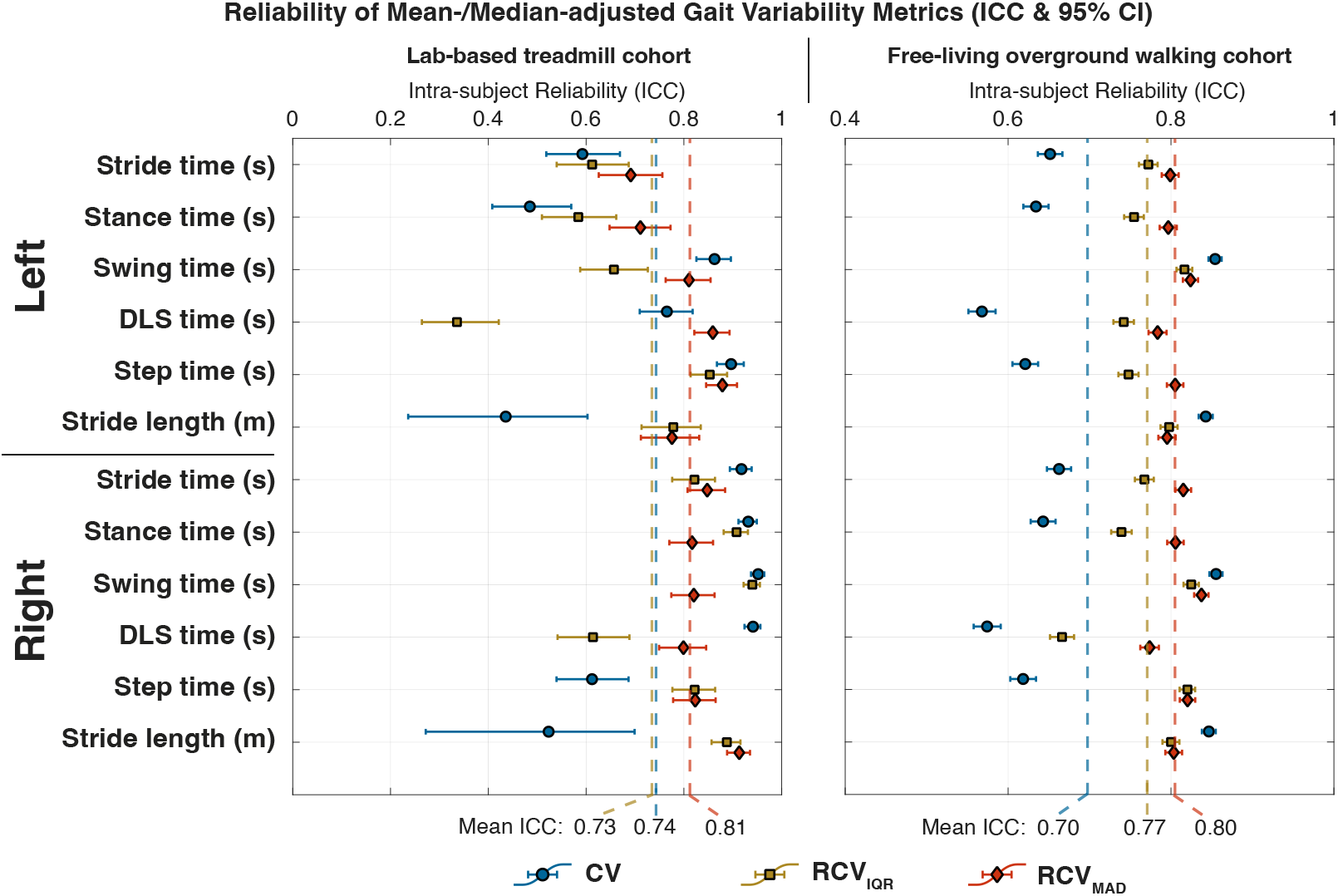
Grouped error bar plots comparing the intra-subject reliability of the conventional Coefficient of Variation (CV), the Robust CV based on the Interquartile Range (RCV_IQR_), and the Robust CV based on the Median Absolute Deviation (RCV_MAD_). Reliability is quantified by the Intra-class Correlation Coefficient (ICC(A,1)), with the shapes (diamonds, squares and circles) representing the mean ICC value and the error bars representing the 95% Confidence Interval (CI), calculated from ten 2-minute moving windows for each participant. Higher ICC values indicate higher reliability. Data are shown for twelve gait parameters (left and right limbs) for two distinct cohorts: (left) lab-based treadmill cohort and (right) free-living overground walking cohort. Dashed vertical lines represent the mean ICC for each metric across all 12 parameters. The RCV_MAD_ is the only metric to demonstrate consistently ‘good’ or ‘excellent’ reliability, while CV and RCV_IQR_ show erratic performance with reliability often in ‘poor’ or ‘moderate’ ranges.

**Fig. 7.**
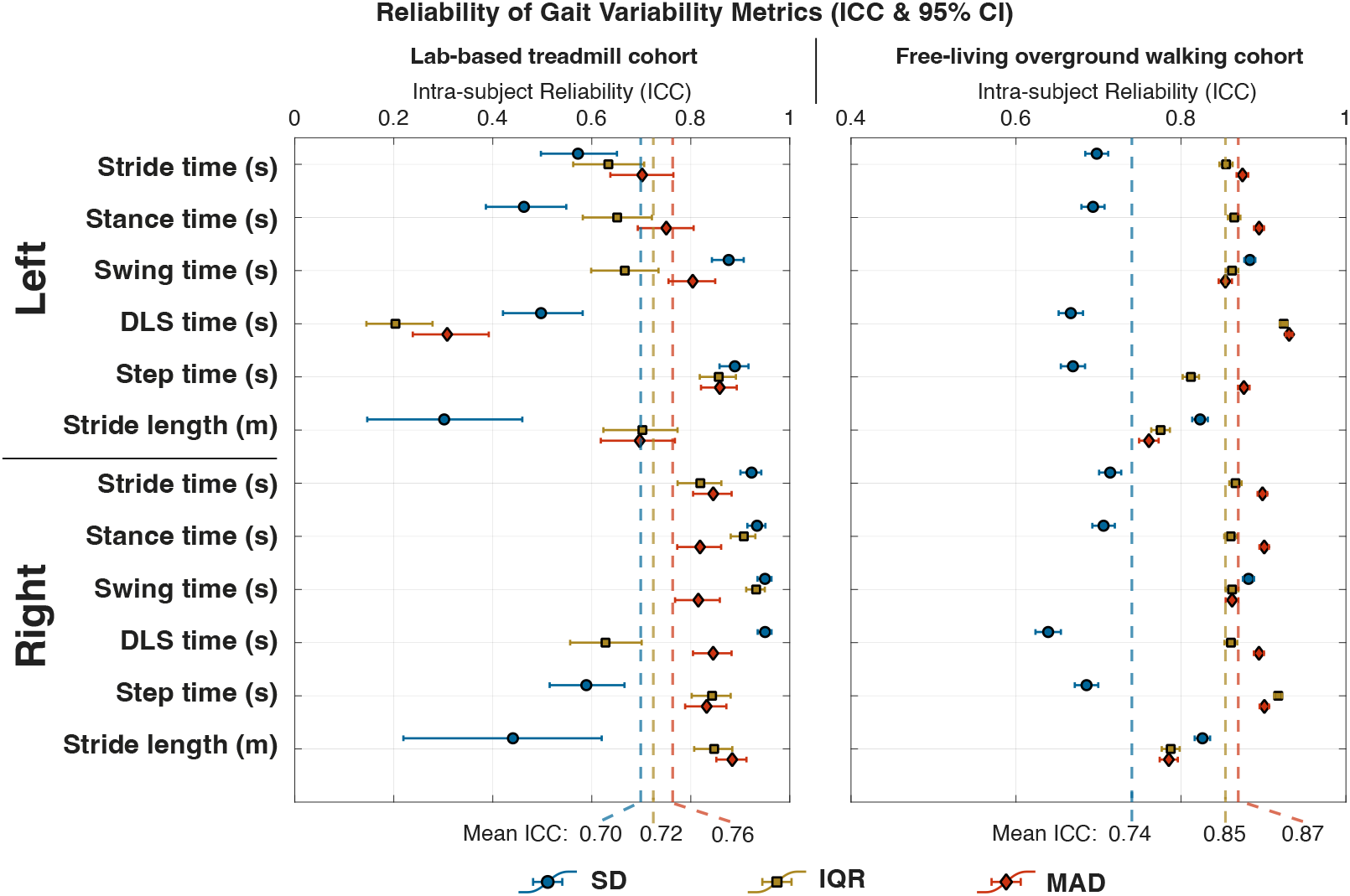
Grouped error bar plots comparing the intra-subject reliability of the Standard Deviation (SD), the Interquartile Range (IQR), and the Median Absolute Deviation (MAD). Reliability is quantified by the Intra-class Correlation Coefficient (ICC(A,1)), with the point representing the ICC value and the error bars representing the 95% Confidence Interval (CI), calculated from ten 2-minute moving windows for each participant. Higher ICC values indicate higher reliability. Data are shown for twelve gait parameters (left and right limbs) for two distinct cohorts: (left) lab-based treadmill cohort and (right) free-living overground walking cohort. Dashed vertical lines represent the mean ICC for each metric across all 12 parameters. The MAD is the only metric to demonstrate consistently ‘good’ or ‘excellent’ reliability, while SD and IQR show erratic performance, with reliability for several parameters falling into the ‘poor’ range.

#### 2.5.2.1 *Reliability of* RCV_MAD_, RCV_IQR_ *and CV*

As shown in Table A4 (Appendix A) and Figure 6, the conventional CV showed highly inconsistent reliability, particularly in the lab-based treadmill cohort. Its reliability ranged from ‘poor’ (e.g. Left Stance Time, ICC = 0.48[0.41–0.57] and Left Stride Time, ICC = 0.44[0.24–0.60]) to ‘excellent’ for some parameters (e.g. Right Swing Time, ICC = 0.95[0.94–0.96]). The RCV_IQR_ reported similar inconsistent reliability, displaying ‘poor’ reliability for Left Step Time (ICC = 0.34[0.26–0.42]) and ‘excellent’ reliability for Right Swing Time (ICC = 0.94[0.92–0.96]). In contrast, the RCV_MAD_ provided the most consistent performance, never achieving ‘poor’ reliability for any of the parameters. In fact, 10 out of the 12 parameters achieved ‘good’ to ‘excellent’ reliability and its lowest value (Left Stride Time, ICC = 0.69) still in the ‘moderate’ range. Overall, the mean ICC of all parameters was the highest for RCV_MAD_, at 0.81 instead of 0.73 and 0.74 for CV and RCV_IQR_, respectively.

In the overground cohort, the results of CV remained mixed, with the ICC of Left Step Time falling in ‘moderate’ range at 0.57[0.55 − 0.58]. The RCV_MAD_ again demonstrated the persistent reliability, with all 12 parameters falling in the ‘good’ to ‘excellent’ range. The superior consistency was reflected with a mean ICC of 0.8, which was again the highest among the three metrics, compared to CV (0.70) and RCV_IQR_ (0.77).

#### 2.5.2.2 Reliability of SD, IQR and MAD

A similar pattern was observed for the absolute dispersion metrics (Table A5 in Appendix A, Figure 7). In the treadmill cohort, the conventional SD showed inconsistent reliability, with ‘poor’ reliability for Left Stride Length (ICC = 0.30[0.15–0.46]) and Left Stance Time (ICC = 0.46[0.39–0.55]). The IQR also performed poorly for Left Step Time (ICC = 0.20[0.14–0.28]). The MAD was the most consistent, with all 12 parameters achieving ‘good’ to ‘excellent’ reliability, with a mean ICC of 0.76.

In the overground cohort, the superiority of the MAD was even more pronounced, achieving highest reliability for 9 of the 12 parameters among the three variability metrics. The mean ICC for MAD in the overground cohort was the highest of any metric across all conditions, underscoring its exceptional reliability for real-world data. Both SD and IQR showed good reliability, but MAD was clearly the most reliable choice.

## 3 Discussion

Gait variability quantifies inter-step fluctuations and captures subtle changes in walking behaviour. Wearable sensors allow characterisation of gait in the real-world under ecological settings. However, conventional metrics of variability such as SD and CV are highly sensitive outliers and therefore poor at assessing intrinsic fluctuations. In this study, we firstly establish normative trajectories and reference values for real-world gait variability across the older adult lifespan, using robust and validated statistical approaches across different walking conditions. The implementation of robust non-parametric measure like RCV_MAD_ allowed us to filter out environmental noise and walking transients, effectively extracts the intrinsic variability of the gait cycle. Leveraging GAMLSS modeling, we demonstrated that movement rhythmicity declines drastically in later life, serving as a sensitive, ecologically valid phenotype of biological aging. Furthermore, we showed that these robust metrics exhibit consistently higher sensitivity in distinguishing older adults with a history of falls compared to conventional metrics, presenting robust metrics as a potential tool for long-term unsupervised monitoring of functional decline in the community.

The age-related increase in robust gait variability likely reflects the decline of central neuromuscular control. While healthy gait is largely an automated motor task from the Central Pattern Generator, ageing leads to a deterioration of this automaticity, resulting a potentially significant increased in reliance on executive function and higher-level cognitive input to regulate gait timing [26]. Recent brain imaging evidence further demonstrated that increased stride length variability correlates with reduced gray matter volume in the hippocampus and supramarginal gyrus, while step time variability maps to the supplementary motor area [6]. In the real world, conventional gait variability measures are affected by the distribution of walking behaviour, contaminated by non-stationary transients such as turns and pauses. By utilising robust metrics that cut through this extrinsic noise, our normative trajectories unveils *the* true underlying physiological deficits. This allows clinicians to distinguish the normative decline of ageing from pathological deviations in stride length and swing time variability, which are potential early signs of cognitive impairment [6].

As shown in the technical validation, the RCV_MAD_ and MAD provide a significantly more consistent and reliable measurement of gait variability than the conventional CV and SD. The demonstrated inconsistency of the conventional CV is a direct consequence of the underlying data distribution reported in our results. Standard deviation is a parametric measure that assumes a normal distribution. However, our analysis revealed that real-world walking data is often highly skewed and heavy-tailed, due to the common occurrence of outliers (e.g. small undetected turns, pauses and obstacle avoidance). The standard deviation involves squaring differences from the mean, which makes it sensitive towards extreme values in real-world data. In contrast, the MAD is a rank-based statistic centered on the median, which works without the normality assumption. It effectively ignores these asymmetric outliers, more suited towards capturing inherent and underlying variability even when substantial proportion of the data is non-normal (skewed or heavy tailed), as seen in our real-world data (Table 1). Hence, RCV_MAD_ and MAD provide a better estimation of dispersion, therefore a more rigorous representation of an individual’s intercycle variability and baseline neuromotor control in real world.

The empirical findings in free-living environments align with previous theoretical work investigating the statistical properties of gait variability metrics [17]. While Chau et al. demonstrated — using clinical lab-based data — that standard deviation is highly vulnerable to outliers, our study validates that this inconsistency is exacerbated in the context of free-living gait and unsupervised monitoring of gait bouts. We show that the weaknesses they identified are further magnified when applied to non-controlled and often ‘messy’ reality of free-living gait data. In line with heart rate variability research studies that have adopted robust metrics e.g. Root Mean Square of Successive Differences (RMSSD) to account for ectopic beats [2], our results indicate that real-world gait analysis should adopt robust statistics like RCV_MAD_ and MAD to account for environmental navigation disturbances.

The clinical implications of this improved consistency are twofold. First, a consistent metric is essential for monitoring long-term, free-living gait. The high consistency of RCV_MAD_ and MAD ensures that an observed change in a patient’s gait variability represents a true physiological change, making it a more consistent measure for intervention outcomes [27]. Second, the standard CV and SD could lead to clinical misinterpretation. An individual’s gait variability might appear to suddenly increase simply because their free-living/real-world assessment included more turns or pauses, potentially leading to an incorrect classification of their fall risk. The RCV_MAD_ and MAD provide a more robust and accurate assessment in these scenarios. In this study, our long-term monitoring case study visually reinforced the statistical findings. The ‘noise’ in the map generated using CV suggest the role of environmental disturbances rather than intra-participant variability. By ignoring these artifacts, the RCV_MAD_ provides a more plausible gait variability analysis in unsupervised free-living gait, accurately identifying genuine, persistent changes in gait behavior rather than transient and momentary outliers.

Our results showed that RCV_MAD_ identified more statistically significant differences between the fallers and non fallers. It consistently produced larger effect sizes for gait parameters in comparison to conventional CV. This suggests RCV_MAD_ has better sensitivity for detecting fall-related changes in gait. However, the effect sizes for all metrics were generally small, indicating that the relationship between gait variability and fall history is probably multifactorial. This finding, derived from a community-dwelling cohort rather than a curated case-control study, strengthens its generalizability and aligns with other studies that have failed to find large effect sizes for gait variability alone [21, 28–30]. This suggests that non-linear approaches, regression or machine learning models would be necessary to effectively classify fall risk [31–33].

This study has several limitations. First, we used two different cohorts in two different settings, which limits our ability to analyze the intra-person effects of switching environments. However, this is partially mitigated by our long-term monitoring dataset, which captures intra-individual variations across varied free-living situations. Future work should assess the same individuals in both settings. Second, we did not assess inter-day reliability. Finally, our cohort consisted of older adults with no major neurological conditions; however, the advantages of RCV in mitigating outlier data may be even more pronounced in clinical populations known for having inherently more variable gait such as patients with Parkinson’s Disease [23, 34]. Future work should focus on using RCV_MAD_ to measure intervention outcomes and establishing its Minimal Clinically Important Difference (MCID).

In conclusion, our findings strongly advocate for the adoption of robust statistical methods, like the RCV_MAD_ and MAD, for processing real-world movement data collected using IMU sensors. This principle extends beyond gait analysis to any domain where sensor data is subject to environmental noise. For gait variability, the RCV_MAD_ and MAD provide a consistent, sensitive, and clinically meaningful metric. Based on our findings we recommend the adoption of robust (RCV_MAD_ and MAD) rather than the conventional variability metrics (CV and SD), particularly in combination with digital wearables. This will significantly improve the accuracy and reliability of gait variability as biomarkers for functional health status that is agnostic to the environment. The robust gait variability outcomes are computationally lean and can easily be implemented in wide variety of digital tools such as smart watches, goggles and other wearables, but also in hearing aids that comprise of IMU allowing rapid uptake.

## 4 Methods

### 4.1 Study participants and data collection protocol

This study utilised data from two different cohorts to ensure a comprehensive analysis across different walking environments and populations. Table 3 summarizes the demographics of the two cohorts.

**Table 3.** Demographics of the lab-based treadmill cohort and the free-living overground walking cohort.

| Characteristic | Treadmill Group |  | Free-living Group | p-value |
| --- | --- | --- | --- | --- |
|  | Young (n=50) | Old (n=50) | (n=2193) |  |
| Age (years), M(SD) | 23.8 (2.3) | 68.8 (5.7) | 74.5 (8.2) | <0.001*** |
| BMI (kg/m<sup>2</sup>), M(SD) | 22.5 (2.9) | 24.0 (4.2) | 24.6 (9.2) | 0.297 |
| Sex (Female), n (%) | 30 (60.0%) | 29 (58.0%) | 921 (42.0%) | 0.996 |

*Note:* p-values for continuous variables (Age, BMI) were calculated using Welch’s t-test. p-value for the categorical variable (Sex) was calculated using the Chi-squared test. Significance levels are denoted as: \*p < 0.05, \*\*p < 0.01, \*\*\*p < 0.001.

#### 4.1.1 Lab-based treadmill cohort

Data were collected from 100 participants recruited from the local community in Zürich, Switzerland. The cohort comprised of two age groups: 50 young adults (23.8 ± 2.3 years, range: 19 to 29 years) and 50 older adults (68.8 ± 5.7 years, range: 60 to 83 years). Inclusion criteria was individuals with no known neurological disorders, movement dysfunction, or fall history. Each participant walked continuously on a treadmill for six minutes at a constant, self-selected comfortable speed. This study was approved by the ETH Zürich Ethics Commission (EK 2020-N-78) and conducted in accordance with the Declaration of Helsinki. All participants provided written informed consent prior to participation in this study.

#### 4.1.2 Free-living overground walking cohort

Data were collected from 2,193 older adults (74.5 ± 8.2 years, range: 60 to 104 years) as part of the Targeted Assessment and Recruitment of Geriatrics for Effective fall prevention Treatments (TARGET) cohort study for assessment of falls and fracture risk in community-dwelling older adults in Singapore. The study is a cross-sectional investigation with stratified random sampling of older adults across Singapore. The study was conducted in the participant’s own home environment. Participants were instructed to walk continuously for at least five minutes, at home in their preferred setting. They were free to turn as needed, and a study interviewer closely followed behind for safety. This study was approved by the National University of Singapore Institutional Review Board (NUS-IRB-2021-168) and ETH Zürich Ethics Commission (EK-2021-N-145), conducted in accordance with the Declaration of Helsinki. All participants provided written informed consent prior to participation in this study.

### 4.2 Data acquisition

Gait data were recorded on the two independent cohorts using wearable Inertial Measurement Unit (IMU) sensors (ZurichMove, Zürich, Switzerland [7, 35]). Sensors were placed on both feet and on the pelvis. Each IMU captured tri-axial acceleration and tri-axial angular velocity with a dynamic range of ±16 g and ±2000°/s, respectively. Data were sampled at 200 Hz and time-synchronised across all sensors.

### 4.3 Data Processing and Gait Parameter Extraction

The raw tri-axial accelerometer and gyroscope signals from each walk test were first filtered to remove noise. A previously validated gait event detection algorithm was then applied to the sensor data from the feet to identify the timing of key gait events, specifically heel-strikes and toe-offs, throughout the entire walking duration [35]. To ensure the variability metrics were not confounded by non-stationary movements, validated turn and pause detection algorithms were also employed [7]. Pauses greater than 5 seconds and continuous turns that are more than 45^*°*^ were removed. This ensured analysis is only carried out on continuous straight-line walking phases, hence the resulting metrics to reflect near steady-state gait variability, especially during free-living overground gait.

From these gait events, a set of spatio-temporal parameters was computed for each gait cycle. Temporal parameters, including stride time, swing time, stance time, step time, and double limb support time, were calculated directly from the event timings [7]. Spatial parameters, such as stride length and gait speed, were estimated using strapdown integration approach on the accelerometer signals from the feet sensors, with orientation estimated using Versatile Quaternion-based Filter (VQF) [36].

### 4.4 Variability and statistical analyses

### 4.5 Variability metrics

Two categories of gait variability metrics were evaluated: absolute measures of dispersion and relative, mean/median-adjusted measures of variability.

Absolute variability was assessed with the SD, the IQR, and the MAD. While SD is widely used to assess gait variability, we included the IQR and MAD as they are well-established robust statistical alternatives that are less sensitive to outliers [37].

For a given sample of gait parameter *x* of *N* data points, these metrics are defined as:

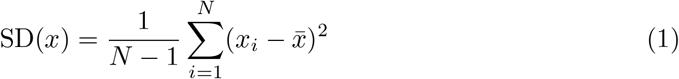

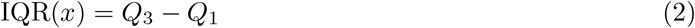

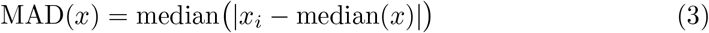

where 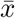 is the sample mean, and *Q*_1_ and *Q*_3_ are the 25th and 75th percentiles of the sample respectively.

For relative variability, we assessed the conventional CV, also a standard variability metric in gait analysis [5, 20]. Alongside CV, we introduced two median-adjusted robust counterparts: the RCV_IQR_ and the RCV_MAD_ [11]. For a given sample of gait parameter *x* of *N* data points, these metrics are defined as:

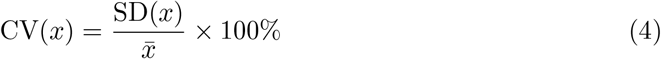

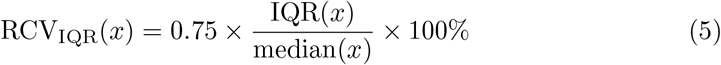

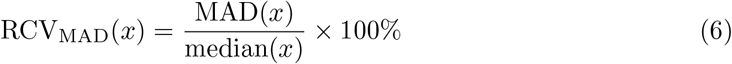

The consistency and reliability analyses were conducted in two groups: the absolute metrics (SD, MAD, and IQR) were compared against each other, and the mean-/median-adjusted relative metrics (CV, RCV_IQR_, and RCV_MAD_) were compared against each other.

#### 4.5.1 Moving window

To assess the intra-subject consistency of the gait variability metrics, a moving window analysis was performed on the continuous time series of the spatio-temporal gait parameters for each participant. Ten overlapping 2-minute windows were extracted from the total walking duration, with the start time of each consecutive window spaced equidistant to the other. This 2-minute duration was selected to ensure robust measurement reliability, as previous research has established that a minimum of 50 gait cycles is required for a reliable gait variability assessment [38]. A 2-minute window comfortably exceeds this threshold for participants with varying gait speed across the cohort, ensuring each window was sufficient for independent and valid estimate of variability.

Within each of the 10 windows, each variability measures was evalutated for each of the seven gait parameters (stride time, swing time, stance time, step time, double limb support time, stride length, and gait speed). This procedure yielded a series of 10 SD, IQR, MAD, CV, RCV_IQR_ and RCV_MAD_ values for each gait parameter for every participant, representing the fluctuation of their gait variability over the course of the walk.

#### 4.5.2 Statistical analysis

The consistency of each variability metric for an individual was quantified by evaluating MAD of the 10 windowed values. MAD, a non-parametric estimator is well-suited for small, likely non-normal data samples [37]. This resulted in a summary metric, for each gait variability measure of each gait parameter for each participant. Smaller MAD values indicate less fluctuation, while larger ICC values indicate high levels of reliability.

To determine if there was a statistically significant difference in consistency between the two metrics, Wilcoxon Signed-Rank Tests were used to compare the paired SD-IQR and SD-MAD values across all participants within each cohort. Similarly, for the relative variability measure, the test was used to compare the paired CV-RCV_IQR_ and CV-RCV_MAD_ values across all participants within each cohort. This non-parametric test was chosen as it is suitable for paired data without assuming a normal distribution. The statistical significance alpha level was set at p *<* 0.05. To quantify the magnitude of the differences between the metrics, effect sizes (*r*) were _calculated as 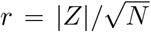_, where *Z* is the standardized test statistic and *N* is the total number of observations.

To evaluate the reliability of each variability metric across the duration of walking bouts, Intraclass Correlation Coefficients (ICC) were calculated. Specifically, a two-way random-effects model selection for absolute agreement based on a single-measurement (ICC(A,1)) was used. For each participant and gait parameter, the ICC was computed using the variability metric values derived from the ten consecutive 2-minute moving windows. Along with the ICC estimate, 95% confidence intervals (CI) were calculated. The analysis was performed in MATLAB (R2025b, The MathWorks, Inc., Natick, MA) using the ‘Intraclass Correlation Coefficient (ICC)’ script [39]. The interpretation of ICC values followed established guidelines [25]: values less than 0.50 indicated ‘poor’ reliability, between 0.50 and 0.75 indicated ‘moderate’, between 0.75 and 0.90 indicated ‘good’, and values greater than 0.90 indicated ‘excellent’ reliability.

### 4.6 Normative Modeling and Statistical Quantification of Aging

To characterise the biological trajectory of gait variability across the aging years and establish normative reference standards, we employed a two-stage statistical framework designed to handle the heavy-tailed, non-Gaussian distributions inherent in real-world mobility data. Normative reference charts of sex-specific centile curves for each robust variability parameter were constructed using Generalised Additive Models for Location, Scale, and Shape (GAMLSS) [40]. Unlike traditional regression which assumes a constant variance and normal distribution, GAMLSS allows for the modeling of distribution parameters i.e. location (*µ*), scale (*σ*), and shape (*υ*) as functions of explanatory variables. Given the strictly positive and skewed nature of variability metrics, we fitted the data to the Box-Cox Cole and Green (BCCG) distribution. The influence of age on the distribution parameters was modeled using penalised B-splines to capture non-linear biological trajectories without overfitting. Separate models were fitted for males and females to generate the 3rd, 15th, 50th (median), 85th, and 97^th^ percentile reference curves (performed using the gamlss package in R, version 5.5-0 [40]).

To statistically quantify the magnitude of age-related decline and formally test for sex differences, we utilised robust quantile regression. While GAMLSS provided the visual reference charts, quantile regression (at the median, *τ* = 0.5) provided the hypothesis testing framework required for non-normal data. For each gait parameter, we fitted a model including Age, Sex, and an Age *×* Sex interaction term. The interaction term was used to empirically test the hypothesis that the rate of aging differs between sexes. The “rate of aging” was quantified as the estimated percentage change in the median variability metric per 10 years of life, with 95% confidence intervals derived using a robust bootstrap method (performed using the quantreg package in R, version 6.1 [41]). Statistical significance was defined as *α <* 0.05.

### 4.7 Assessing clinical utility of variability measure: fall risk

To evaluate the clinical utility of variability metrics, their ability to differentiate between fallers and non-fallers was investigated using data from the free-living overground walking cohort. Participants were categorised into these two groups based on their self-reported answer (“Yes” or “No”) to the question “Have you fallen even once in the past 12 months?”. Statistical differences between the two groups were assessed using the non-parametric Mann-Whitney U-test, p-values were reported for statistical significance. To illustrate the effect size of the difference between the two groups, Cohen’s d was calculated for each parameter.

### 4.8 Assessing gait variability spatially in unsupervised monitoring data

Unsupervised longitudinal movement monitoring data is collected as part of the ongoing Built Environment in Falls and Arthritis (BE-FIT) study which aimed to understand the neighborhood built environment influence on the mobility, participation, and psychosocial health of older adults with knee osteoarthritis and/or falls. The protocol included a 28-day unsupervised movement monitoring phase which intended to capture daily movement behaviors and functional activities in free-living conditions using wearable sensors. All participants provided written informed consent, and the study procedures were approved by the local institutional review board in accordance with the Declaration of Helsinki.

To capture continuous kinematic data, participants were instrumented with a tri-axial IMU sensor Axivity AX6 (Axivity Ltd) secured to the L5 vertebra using waterproof tapes. The device was configured to record acceleration and angular velocity with a dynamic range of ±8 g and ±1000°/s, respectively. Data were sampled at 100 Hz. To provide environmental context for these gait patterns, participants were requested to carry a smartphone (iPhone 14) running a custom GPS application Sensor Logger to collect geospatial coordinates and altitude, and with timestamps.

Raw IMU data were processed using the SciKit Digital Health (SKDH) Python framework [42]. In particular, activity classification was first performed to detect walking activities. Using a pre-trained Light Gradient Boosting Machine (LGBM) model non-overlapping 3-second windows of acceleration data are classified as either gait or non-gait. Fragmented gait bouts separated by less than 60 seconds were merged into single bout. Gait bouts with a total duration of less than 5 minutes were excluded. Within these sustained bouts, gait events (i.e. initial contact and final contact) were detected from the lumbar acceleration signal using Continuous Wavelet Transform (CWT) methods [43], provided in the SKDH pipeline. These events enabled the computation of spatio-temporal parameters, including stride time, stance time, swing time, double limb support time, step time, and stride length [44].

The extracted gait bouts were synchronised with GPS data using timestamps. Bouts occurring within the 50 meters of the participants’ home were filtered out. The final dataset consisted of 505 five-minute gait segments obtained from 9 participants. For each segment, gait variability measures i.e. CV, RCV_IQR_, and the RCV_MAD_ were calculated for six gait parameters. Consistency of the gait variability metrics were computed on 10 overlapping 2-min windows, similar to the Section 4.4.

## Supplementary information

## Acknowledgements

The research was conducted at the Future Health Technologies at the Singapore-ETH Centre, which was established collaboratively between ETH Zürich and the National Research Foundation Singapore. This research is supported by the National Research Foundation Singapore (NRF) under its Campus for Research Excellence and Technological Enterprise (CREATE) programme. The data collection for the lab-based treadmill cohort and the corresponding research positions were funded by Innosuisse (Swiss Innovation Agency), Grant Number 120.497 IP-LS.

## Author Contributions

K.T. conceptualized the work. The study design and data acquisition were conducted by sub-teams specific to each cohort: Y.K, A.F., M.G., N.S. for the lab-based study; K.T., K.T., V.K., R.M., A.C., D.M. and N.S. for the overground free-living study; and K.F. and N.S. for the long-term monitoring case study. K.T. and K.F. performed the formal data analysis, coding, visualization, and interpretation across all datasets. K.T. prepared the initial draft of the manuscript. All authors actively participated in the comprehensive review, refinement, and finalization of the manuscript. N.S. supervised the project. All authors have read and agreed to the published version of the manuscript.

## Ethics Declarations

This lab-based treadmill cohort study was approved by the ETH Zürich Ethics Commission (EK 2020-N-78). The free-living overground walking cohort study was approved by the National University of Singapore Institutional Review Board (NUS-IRB-2021-168) and ETH Zürich Ethics Commission (EK-2021-N-145). Both studies were conducted in accordance with the Declaration of Helsinki. All participants provided written informed consent prior to participation in this study.

## Data availability

The wearable sensor gait data from both lab-based treadmill cohort and free-living overground walking cohort are not publicly available. Request for access to the lab based treadmill cohort dataset can directed to.

## Code availability

The analyses were implemented in MATLAB 2025b and the code is available under the MIT license in the following GitLab repository: https://gitlab.ethz.ch/kaitan/robust-cv-comparison.

## Appendix A Supplementary materials

**Table A1.** Robust quantile regression analysis quantifying the effects of age, sex, and their interaction on ambulatory variability parameters.

| Parameter<br>(Variability) | Significance levels |  |  | Percentage point increase per 10 years |  |
| --- | --- | --- | --- | --- | --- |
|  | Age | Sex | Interaction | Female | Male |
| **Left** |  |  |  |  |  |
| Stride Time | <0.001 *** | 0.788 | 0.465 | 0.36 [0.19 – 0.53] | 0.25 [0.01 – 0.49] |
| Stance Time | 0.001 ** | 0.923 | 0.736 | 0.42 [0.16 – 0.69] | 0.35 [0.03 – 0.68] |
| Swing Time | <0.001 *** | 0.901 | 0.732 | 0.78 [0.54 – 1.02] | 0.72 [0.47 – 0.98] |
| Step Time | 0.001 ** | 0.497 | 0.306 | 0.50 [0.20 – 0.81] | 0.30 [0.04 – 0.55] |
| DLS Time | 0.346 | 0.639 | 0.501 | 0.28 [–0.31 – 0.88] | –0.00 [–0.62 – 0.62] |
| Stride Length | <0.001 *** | 0.522 | 0.557 | 1.29 [0.94 – 1.63] | 1.45 [1.04 – 1.86] |
| **Right** |  |  |  |  |  |
| Stride Time | 0.002 ** | 0.634 | 0.325 | 0.35 [0.13 – 0.57] | 0.21 [0.01 – 0.40] |
| Stance Time | <0.001 *** | 0.515 | 0.251 | 0.62 [0.29 – 0.96] | 0.36 [0.03 – 0.69] |
| Swing Time | <0.001 *** | 0.185 | 0.144 | 0.90 [0.65 – 1.14] | 0.60 [0.31 – 0.89] |
| Step Time | <0.001 *** | 0.351 | 0.181 | 0.53 [0.23 – 0.83] | 0.24 [–0.04 – 0.53] |
| DLS Time | 0.103 | 0.857 | 0.576 | 0.45 [–0.11 – 1.01] | 0.20 [–0.47 – 0.86] |
| Stride Length | <0.001 *** | 0.985 | 0.950 | 1.20 [0.88 – 1.51] | 1.22 [0.80 – 1.64] |

Note: Significance levels derived from robust quantile regression models ( $N = 2, 193$ ).
Decadal estimates represent the median percentage point increase with bootstrap 95% confidence intervals.
DLS - Double Limb Support.
\* $p < 0.05$ , \*\* $p < 0.01$ , \*\*\* $p < 0.001$ .

**Fig. A1.**
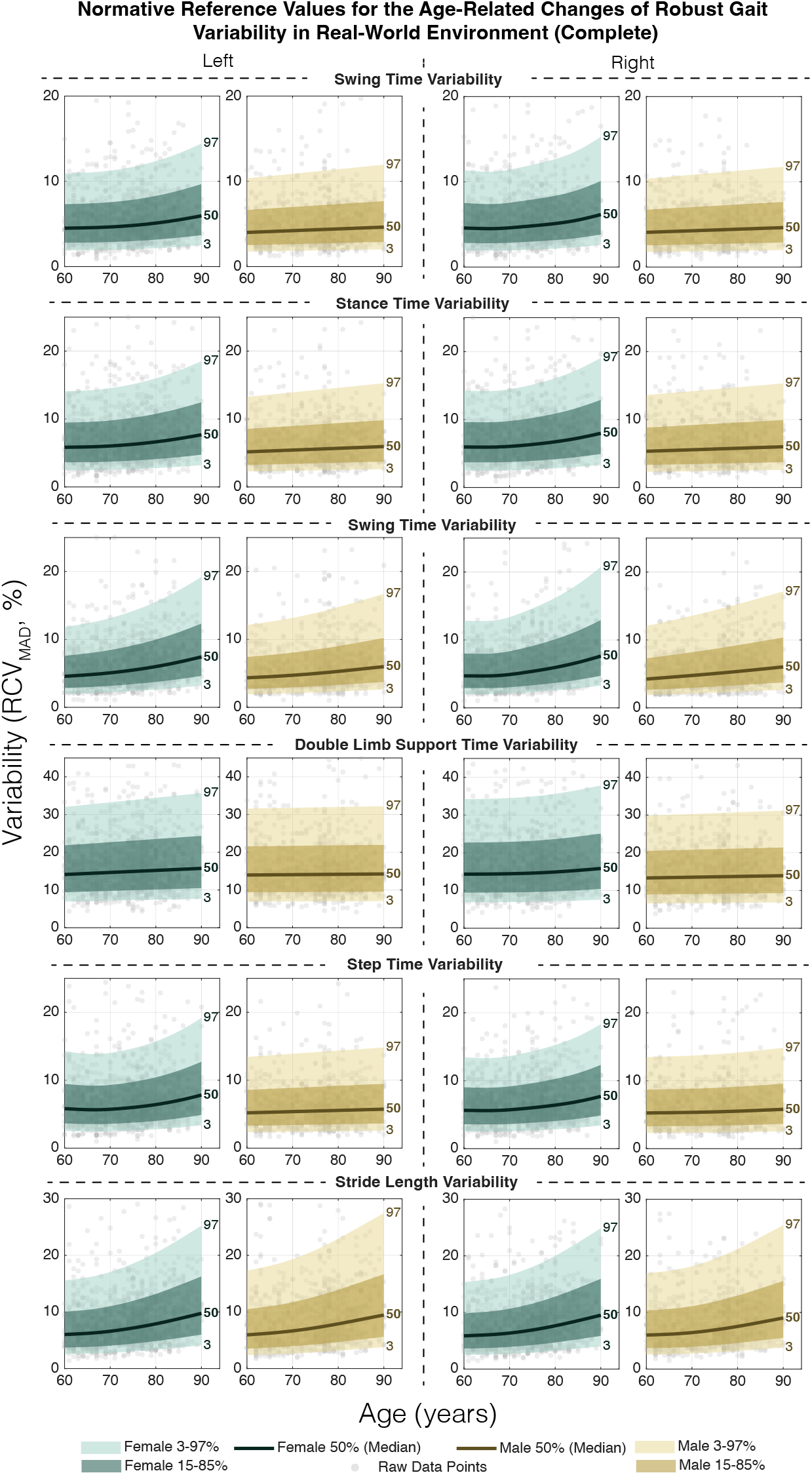
Age-related trajectories of robust gait variability (RCV_MAD_) in free-living environments: Generalised Additive Models for Location, Scale, and Shape (GAMLSS) centile charts (3rd, 15th, 50th, 85th, and 97th percentiles) depicting the normative changes of all gait parameters across the older adult lifespan (60–90 years), stratified by sex (Females: Green; Males: Yellow). The median (50th percentile) trajectory highlights a progressive loss of movement spatio-temporal rhythmicity, identifying robust variability as a sensitive digital biomarker for monitoring age-related functional decline.

**Table A2.** Comparison the consistency of CV, RCV_IQR_ and RCV_IQR_ in estimating gait parameter variability.

| Gait Parameter | Metric | Lab-based treadmill data |  |  | Overground walking data |  |  |
| --- | --- | --- | --- | --- | --- | --- | --- |
|  |  | Median Consistency (IQR) [10<sup>-2</sup>] | Effect size | p-value | Median Consistency (IQR) [10<sup>-2</sup>] | Effect size | p-value |
| Left |  |  |  |  |  |  |  |
| Stride time | CV | 7.30 (4.37-14.49) | - | - | 92.92 (40.87-243.44) | - | - |
|  | RCV<sub>IQR</sub> | 7.81 (0.66-17.46) | 0.13 | 0.10 | 66.33 (39.53-115.54) | 0.36 | < 0.001 |
|  | RCV<sub>MAD</sub> | 0.43 (0.18-3.09) | 0.59 | < 0.001 | 42.82 (25.78-72.35) | 0.65 | < 0.001 |
| Stance time | CV | 9.77 (5.31-19.65) | - | - | 128.59 (55.92-361.70) | - | - |
|  | RCV<sub>IQR</sub> | 3.11 (0.92-17.40) | 0.31 | < 0.001 | 86.73 (50.45-159.58) | 0.39 | < 0.001 |
|  | RCV<sub>MAD</sub> | 0.60 (0.00-1.58) | 0.72 | < 0.001 | 58.60 (33.29-97.35) | 0.67 | < 0.001 |
| Swing time | CV | 9.80 (6.69-19.21) | - | - | 71.84 (39.90-126.14) | - | - |
|  | RCV<sub>IQR</sub> | 4.65 (0.74-31.42) | 0.11 | 0.15 | 72.82 (43.72-122.26) | -0.02 | 0.79 |
|  | RCV<sub>MAD</sub> | 0.58 (0.00-2.57) | 0.58 | < 0.001 | 49.62 (19.87-87.31) | 0.44 | < 0.001 |
| Double limb support time | CV | 45.81 (27.92-83.11) | - | - | 275.83 (127.14-729.09) | - | - |
|  | RCV<sub>IQR</sub> | 31.25 (0.00-168.56) | -0.04 | 0.65 | 221.71 (124.21-384.74) | 0.29 | < 0.001 |
|  | RCV<sub>MAD</sub> | 0.00 (0.00-20.50) | 0.64 | < 0.001 | 125.00 (56.83-221.77) | 0.63 | < 0.001 |
| Step time | CV | 11.44 (6.15-22.72) | - | - | 94.13 (46.79-227.88) | - | - |
|  | RCV<sub>IQR</sub> | 5.65 (0.76-32.17) | 0.14 | 0.08 | 78.45 (47.84-135.64) | 0.27 | < 0.001 |
|  | RCV<sub>MAD</sub> | 0.78 (0.00-1.75) | 0.63 | < 0.001 | 53.71 (29.25-88.39) | 0.58 | < 0.001 |
| Stride Length | CV | 34.96 (12.48-117.74) | - | - | 121.46 (66.14-219.06) | - | - |
|  | RCV<sub>IQR</sub> | 36.29 (16.69-89.41) | -0.14 | 0.92 | 109.56 (61.68-194.75) | 0.03 | 0.05 |
|  | RCV<sub>MAD</sub> | 23.85 (11.97-61.96) | 0.55 | < 0.001 | 68.48 (39.34-117.70) | 0.51 | < 0.001 |
| Right |  |  |  |  |  |  |  |
| Stride time | CV | 8.53 (4.76-12.94) | - | - | 90.25 (40.90-237.73) | - | - |
|  | RCV<sub>IQR</sub> | 4.47 (0.47-20.33) | 0.10 | 0.16 | 67.03 (38.20-118.52) | 0.33 | < 0.001 |
|  | RCV<sub>MAD</sub> | 0.57 (0.18-5.54) | 0.53 | < 0.001 | 42.81 (25.35-73.20) | 0.63 | < 0.001 |
| Stance time | CV | 9.66 (6.33-16.11) | - | - | 124.75 (55.29-345.75) | - | - |
|  | RCV<sub>IQR</sub> | 3.95 (0.52-25.26) | 0.11 | 0.14 | 87.88 (50.05-159.13) | 0.36 | < 0.001 |
|  | RCV<sub>MAD</sub> | 0.63 (0.00-3.38) | 0.48 | < 0.001 | 57.65 (34.16-98.70) | 0.66 | < 0.001 |
| Swing time | CV | 10.21 (6.89-19.31) | - | - | 69.63 (40.62-120.61) | - | - |
|  | RCV<sub>IQR</sub> | 3.36 (0.00-27.29) | 0.13 | 0.1 | 73.51 (43.28-125.47) | -0.06 | 1 |
|  | RCV<sub>MAD</sub> | 0.56 (0.00-2.58) | 0.49 | < 0.001 | 50.59 (18.66-87.25) | 0.42 | < 0.001 |
| Double limb support time | CV | 66.16 (43.53-106.35) | - | - | 297.69 (132.95-713.81) | - | - |
|  | RCV<sub>IQR</sub> | 34.36 (0.00-129.52) | 0.20 | 0.02 | 223.21 (127.33-395.08) | 0.30 | < 0.001 |
|  | RCV<sub>MAD</sub> | 0.00 (0.00-23.81) | 0.69 | < 0.001 | 131.24 (58.88-225.00) | 0.67 | < 0.001 |
| Step time | CV | 9.94 (6.46-19.11) | - | - | 93.76 (46.34-232.15) | - | - |
|  | RCV<sub>IQR</sub> | 3.74 (0.63-30.59) | 0.24 | 0.01 | 78.78 (46.91-137.60) | 0.27 | < 0.001 |
|  | RCV<sub>MAD</sub> | 0.53 (0.00-1.92) | 0.62 | < 0.001 | 54.23 (30.29-87.61) | 0.58 | < 0.001 |
| Stride Length | CV | 11.92 (7.55-22.80) | - | - | 118.31 (65.06-208.83) | - | - |
|  | RCV<sub>IQR</sub> | 17.62 (9.71-27.36) | -0.18 | 0.96 | 107.13 (59.10-194.86) | 0.03 | 0.09 |
|  | RCV<sub>MAD</sub> | 11.05 (7.52-17.36) | 0.30 | < 0.001 | 66.38 (38.16-117.02) | 0.51 | < 0.001 |

**Table A3.** Comparison the consistency of SD, IQR and MAD in estimating gait parameter variability.

| Gait Parameter | Metric | Lab-based treadmill data |  |  | Overground walking data |  |  |
| --- | --- | --- | --- | --- | --- | --- | --- |
|  |  | Median Consistency (IQR) [10<sup>-3</sup>] | Effect size | p-value | Median Consistency (IQR) [10<sup>-3</sup>] | Effect size | p-value |
| Left |  |  |  |  |  |  |  |
| Stride time | SD | 0.81 (0.49-1.55) | - | - | 14.25 (5.91-38.04) | - | - |
|  | IQR | 0.94 (0.00-1.88) | 0.15 | 0.07 | 9.37 (5.16-17.34) | 0.39 | <0.001 |
|  | MAD | 0.00 (0.00-0.00) | 0.59 | <0.001 | 6.25 (3.75-11.25) | 0.65 | <0.001 |
| Stance time | SD | 0.61 (0.33-1.32) | - | - | 13.33 (5.11-40.17) | - | - |
|  | IQR | 0.00 (0.00-1.41) | 0.37 | <0.001 | 7.50 (4.22-15.47) | 0.44 | <0.001 |
|  | MAD | 0.00 (0.00-0.00) | 0.72 | <0.001 | 5.00 (2.50-10.00) | 0.68 | <0.001 |
| Swing time | SD | 0.50 (0.33-0.98) | - | - | 3.56 (1.99-6.45) | - | - |
|  | IQR | 0.00 (0.00-1.64) | 0.16 | 0.06 | 3.75 (1.88-6.09) | 0.00 | 0.49 |
|  | MAD | 0.00 (0.00-0.00) | 0.61 | <0.001 | 2.50 (0.00-5.00) | 0.44 | <0.001 |
| Double limb support time | SD | 0.40 (0.22-0.54) | - | - | 8.32 (2.88-29.63) | - | - |
|  | IQR | 0.00 (0.00-0.94) | 0.21 | 0.02 | 3.75 (1.88-7.50) | 0.42 | <0.001 |
|  | MAD | 0.00 (0.00-0.00) | 0.69 | <0.001 | 2.50 (0.00-5.00) | 0.68 | <0.001 |
| Step time | SD | 0.62 (0.34-1.29) | - | - | 7.83 (3.22-19.50) | - | - |
|  | IQR | 0.00 (0.00-1.88) | 0.18 | 0.03 | 5.63 (3.28-10.31) | 0.32 | <0.001 |
|  | MAD | 0.00 (0.00-0.00) | 0.61 | <0.001 | 3.75 (2.50-6.25) | 0.59 | <0.001 |
| Stride Length | SD | 3.49 (1.24-10.08) | - | - | 7.58 (4.22-13.37) | - | - |
|  | IQR | 3.92 (1.85-8.03) | -0.18 | 0.97 | 7.44 (4.39-12.89) | -0.04 | 0.96 |
|  | MAD | 2.45 (1.05-5.76) | 0.54 | <0.001 | 4.63 (2.79-7.79) | 0.45 | <0.001 |
| Right |  |  |  |  |  |  |  |
| Stride time | SD | 1.00 (0.52-1.49) | - | - | 14.01 (5.85-38.72) | - | - |
|  | IQR | 0.47 (0.00-2.11) | 0.14 | 0.08 | 9.38 (5.16-17.81) | 0.36 | <0.001 |
|  | MAD | 0.00 (0.00-0.00) | 0.54 | <0.001 | 6.25 (3.75-11.25) | 0.63 | <0.001 |
| Stance time | SD | 0.63 (0.42-1.01) | - | - | 13.27 (4.94-38.53) | - | - |
|  | IQR | 0.00 (0.00-1.87) | 0.16 | 0.06 | 7.50 (4.22-15.94) | 0.40 | <0.001 |
|  | MAD | 0.00 (0.00-0.00) | 0.49 | <0.001 | 5.00 (2.50-10.00) | 0.66 | <0.001 |
| Swing time | SD | 0.54 (0.31-0.97) | - | - | 3.46 (2.00-6.32) | - | - |
|  | IQR | 0.00 (0.00-1.41) | 0.19 | 0.03 | 3.75 (2.34-6.56) | -0.07 | 1 |
|  | MAD | 0.00 (0.00-0.00) | 0.52 | <0.001 | 2.50 (0.00-5.00) | 0.42 | <0.001 |
| Double limb support time | SD | 0.39 (0.25-0.66) | - | - | 8.44 (2.49-21.00) | - | - |
|  | IQR | 0.00 (0.00-0.47) | 0.29 | 0 | 3.75 (1.88-7.50) | 0.43 | <0.001 |
|  | MAD | 0.00 (0.00-0.00) | 0.71 | <0.001 | 2.50 (0.00-5.00) | 0.69 | <0.001 |
| Step time | SD | 0.57 (0.36-1.17) | - | - | 7.20 (3.36-19.06) | - | - |
|  | IQR | 0.00 (0.00-1.88) | 0.27 | 0 | 5.63 (3.28-10.31) | 0.31 | <0.001 |
|  | MAD | 0.00 (0.00-0.00) | 0.63 | <0.001 | 3.75 (2.50-6.25) | 0.61 | <0.001 |
| Stride Length | SD | 1.33 (0.84-2.44) | - | - | 7.44 (4.24-13.39) | - | - |
|  | IQR | 1.86 (1.11-2.73) | -0.17 | 0.95 | 7.30 (4.31-12.88) | -0.03 | 0.9 |
|  | MAD | 1.17 (0.85-1.78) | 0.32 | <0.001 | 4.51 (2.75-7.54) | 0.47 | <0.001 |

**Table A4.** Intra-subject Reliability (ICC; 95% Confidence Interval) of Gait Variability Metrics (Absolute Dispersion)

| Gait Parameter | Treadmill walking ICC (95% Confidence Interval) |  |  | Overground walking ICC (95% Confidence Interval) |  |  |
| --- | --- | --- | --- | --- | --- | --- |
|  | SD | IQR | MAD | SD | IQR | MAD |
| **Left** |  |  |  |  |  |  |
| Stride Time | 0.57 (0.50–0.65) | 0.63 (0.56–0.71) | 0.70 (0.64–0.76) | 0.70 (0.68–0.71) | 0.85 (0.85–0.86) | 0.87 (0.87–0.88) |
| Stance Time | 0.46 (0.39–0.55) | 0.65 (0.58–0.72) | 0.75 (0.69–0.81) | 0.69 (0.68–0.71) | 0.86 (0.86–0.87) | 0.89 (0.89–0.90) |
| Swing Time | 0.88 (0.84–0.91) | 0.67 (0.60–0.73) | 0.80 (0.76–0.85) | 0.88 (0.88–0.89) | 0.86 (0.85–0.87) | 0.85 (0.85–0.86) |
| Step Time | 0.50 (0.42–0.58) | 0.20 (0.14–0.28) | 0.31 (0.24–0.39) | 0.67 (0.65–0.68) | 0.92 (0.92–0.93) | 0.93 (0.93–0.94) |
| DLS Time | 0.89 (0.86–0.92) | 0.86 (0.82–0.89) | 0.86 (0.82–0.89) | 0.67 (0.65–0.68) | 0.81 (0.80–0.82) | 0.88 (0.87–0.88) |
| Stride Length | 0.30 (0.15–0.46) | 0.70 (0.62–0.77) | 0.70 (0.62–0.77) | 0.82 (0.81–0.83) | 0.78 (0.76–0.79) | 0.76 (0.75–0.77) |
| **Right** |  |  |  |  |  |  |
| Stride Time | 0.92 (0.90–0.94) | 0.82 (0.77–0.86) | 0.85 (0.80–0.88) | 0.71 (0.70–0.73) | 0.87 (0.86–0.87) | 0.90 (0.89–0.90) |
| Stance Time | 0.93 (0.91–0.95) | 0.91 (0.88–0.93) | 0.82 (0.77–0.86) | 0.71 (0.69–0.72) | 0.86 (0.85–0.87) | 0.90 (0.90–0.91) |
| Swing Time | 0.95 (0.93–0.96) | 0.93 (0.91–0.95) | 0.82 (0.77–0.86) | 0.88 (0.88–0.89) | 0.86 (0.85–0.87) | 0.86 (0.85–0.87) |
| Step Time | 0.95 (0.94–0.96) | 0.63 (0.56–0.70) | 0.85 (0.80–0.88) | 0.64 (0.62–0.65) | 0.86 (0.85–0.87) | 0.89 (0.89–0.90) |
| DLS Time | 0.59 (0.51–0.67) | 0.84 (0.80–0.88) | 0.83 (0.79–0.87) | 0.69 (0.67–0.70) | 0.92 (0.91–0.92) | 0.90 (0.90–0.91) |
| Stride Length | 0.44 (0.22–0.62) | 0.85 (0.81–0.88) | 0.88 (0.85–0.91) | 0.83 (0.82–0.84) | 0.79 (0.78–0.80) | 0.79 (0.77–0.80) |

**Table A5.** Intra-subject Reliability (ICC; 95% Confidence Interval) of Mean-/Median-Adjusted Gait Variability Metrics.

| Gait Variability | Treadmill walking ICC (95% Confidence Interval) |  |  | Overground walking ICC (95% Confidence Interval) |  |  |
| --- | --- | --- | --- | --- | --- | --- |
|  | CV | RCV<sub>IQR</sub> | RCV<sub>MAD</sub> | CV | RCV<sub>IQR</sub> | RCV<sub>MAD</sub> |
| **Left** |  |  |  |  |  |  |
| Stride Time | 0.59 (0.52–0.67) | 0.61 (0.54–0.69) | 0.69 (0.63–0.76) | 0.65 (0.64–0.67) | 0.77 (0.76–0.78) | 0.80 (0.79–0.81) |
| Stance Time | 0.48 (0.41–0.57) | 0.58 (0.51–0.66) | 0.71 (0.65–0.77) | 0.63 (0.62–0.65) | 0.75 (0.74–0.77) | 0.80 (0.79–0.81) |
| Swing Time | 0.86 (0.83–0.90) | 0.66 (0.59–0.73) | 0.81 (0.76–0.85) | 0.85 (0.85–0.86) | 0.82 (0.81–0.83) | 0.82 (0.81–0.83) |
| Step Time | 0.77 (0.71–0.82) | 0.34 (0.26–0.42) | 0.86 (0.82–0.89) | 0.57 (0.55–0.58) | 0.74 (0.73–0.75) | 0.78 (0.77–0.79) |
| DLS Time | 0.90 (0.87–0.92) | 0.85 (0.81–0.89) | 0.88 (0.85–0.91) | 0.62 (0.61–0.64) | 0.75 (0.74–0.76) | 0.81 (0.79–0.82) |
| Stride Length | 0.44 (0.24–0.60) | 0.78 (0.71–0.83) | 0.78 (0.71–0.83) | 0.84 (0.83–0.85) | 0.80 (0.79–0.81) | 0.80 (0.78–0.81) |
| **Right** |  |  |  |  |  |  |
| Stride Time | 0.92 (0.89–0.94) | 0.82 (0.78–0.86) | 0.85 (0.81–0.88) | 0.66 (0.65–0.68) | 0.77 (0.76–0.78) | 0.82 (0.81–0.82) |
| Stance Time | 0.93 (0.91–0.95) | 0.91 (0.88–0.93) | 0.82 (0.77–0.86) | 0.64 (0.63–0.66) | 0.74 (0.73–0.75) | 0.81 (0.80–0.82) |
| Swing Time | 0.95 (0.94–0.96) | 0.94 (0.92–0.96) | 0.82 (0.77–0.86) | 0.86 (0.85–0.86) | 0.82 (0.82–0.83) | 0.84 (0.83–0.85) |
| Step Time | 0.94 (0.92–0.96) | 0.61 (0.54–0.69) | 0.80 (0.75–0.85) | 0.57 (0.56–0.59) | 0.67 (0.65–0.68) | 0.77 (0.76–0.79) |
| DLS Time | 0.61 (0.54–0.69) | 0.82 (0.78–0.86) | 0.82 (0.78–0.86) | 0.62 (0.60–0.63) | 0.82 (0.81–0.83) | 0.82 (0.81–0.83) |
| Stride Length | 0.52 (0.27–0.70) | 0.89 (0.86–0.92) | 0.91 (0.89–0.94) | 0.85 (0.84–0.85) | 0.80 (0.79–0.81) | 0.80 (0.79–0.81) |

